# Opioid use disorder and brain health: observational and genetic associations

**DOI:** 10.1101/2024.12.18.24319222

**Authors:** Sara Javidnia, James M. Roe, Ville Karhunen, Dipender Gill, Steven Bell, Joseph Deak, Daniel Levey, Toinet Cronje, Stephen Burgess, Joel Gelernter, Klaus P. Ebmeier, Anya Topiwala

## Abstract

**Background:** The long-term impact of opioid use disorder (OUD) on brain health has been little explored although of potentially high public health importance.

**Objectives:** To investigate the potential causal impact of OUD on later life brain health outcomes, including dementia, stroke and brain structure.

**Methods:** Observational and Mendelian randomization (MR) analyses were conducted. Participants included in observational analyses were enrolled in the US Million Veteran Program (MVP). Cox proportional hazards were used to examine the association between electronic health record (EHR)-derived diagnoses of OUD and incident dementia in European and African ancestry populations. Two-sample MR was applied to explore the association between genetic predisposition to OUD and dementia, as well as key endophenotypes including brain structure. Several analyses were used for insight into aetiological pathways, including cis-MR to assess the impact of genetically-proxied opioid receptor perturbation, Bayesian colocalization, and polygenic risk score analyses of longitudinal brain changes in non-opioid users from the Lifebrain project (n=229).

**Results:** Amongst 222,518 MVP participants, 8397 developed dementia during follow up. Participants with OUD (n=9,399) were younger and more likely to be male. In observational analyses, OUD was associated with a higher risk of incident all-cause dementia (hazard ratio [HR]=1.56, 95% confidence interval [CI] [1.39,1.76];p=2.23×10^-13^), Alzheimer’s [HR=1.40[1.04,1.87]; p=0.02) and vascular dementia (HR=1.49[1.19,1.86];p=0.0004). In the genetic analysis, genetically-proxied OUD also associated with higher dementia risk. A doubling in genetically-proxied OUD prevalence was associated with a 77% increase in odds of dementia (IVW OR=1.77[1.43,2.19];p=1.69×10^⁻⁷^). Variation in μ-opioid receptor genes were strongly associated with dementia risk. No significant associations were observed with brain structure in non-opioid users, nor in lower powered non-European ancestry groups.

**Conclusions:** These findings suggest a potential causal impact of opioid use disorder on dementia. Genetic analyses supported an aetiological role of μ-opioid receptor pathways. Further pharmacovigilance and investigation into the long-term effects of opioids on brain health are warranted.

## Introduction

The opioid epidemic is a global public health crisis (Upp et al., 2020). While much attention has been focused on the short-term consequences of opioid use and use disorder (OUD) (Hoffman et al., 2020; Ledger et al., 2020), less is known about their long-term impacts. The latter are difficult to assess in randomized controlled trials because of the necessary duration and recruitment difficulties of this clinical population. Emerging evidence suggests that opioid use may accelerate cognitive decline (Bhatia et al., 2023; Dhingra., 2015). There is an urgent need for rigorous research into the potential long-term neurodegenerative consequences of opioid use disorder, particularly as populations continue to age and the prevalence of age-related cognitive impairment rises.

Several observational studies have reported that individuals with a history of opioid use disorder are at a higher risk of cognitive impairment and neurodegenerative diseases, including dementia (Pourhadi et al., 2024; Qeadan et al., 2023; Levine et al., 2023; Badr et al., 2023; Verdejo-Garcia et al., 2019). Links between opioids and cognitive decline are biologically plausible. Opioids primarily exert their effects through the μ- and δ-opioid receptors (encoded by *OPRM1* and *OPRD1* genes), which influence critical neurobiological processes implicated in dementia (Scuteri et al., 2020; Dublin et al., 2015), such as neurotransmitter release, neuroinflammation, and synaptic plasticity (Xu et al., 2023; Cuitavi et al., 2023). However, previous studies were observational in nature. The potential for residual confounding, by comorbidities often associated with OUD, including chronic pain, psychiatric disorders (Hser et al., 2017), and socio-economic factors (Sulley et al., 2020) is high, as well as reverse causation, making it difficult to establish causality (Neelamegam et al., 2021; Dublin et al., 2015).

Mendelian randomization (MR) (Burgess et al., 2023), a quasi-experimental method, overcomes several limitations of traditional observational methods. It uses genetic variants as instruments to proxy exposures. MR can be extended to offer mechanistic insights, by restricting instrumental genetic variants to single gene regions encoding drug targets (cis variants) (Gill et al., 2021). Here, we triangulate large-scale observational and genetic analyses of opioid use disorder (OUD), a severe phenotype of physiological opioid dependence, and dementia outcomes. We perform several analyses to understand aetiological pathways, including cis-MR, Bayesian colocalization, and polygenic risk score analyses in a non-opioid exposed sample to disentangle effects driven by genetic susceptibility to OUD from those resulting from actual opioid exposure.

## Methods

### Study populations

The observational analyses included two cohorts. First, for analysis of dementia outcomes, participants were drawn from the US Million Veteran Program (MVP), that has been enrolling US veterans (mean age = 61.52 [SD =10.13]) since 2011 (see Supplemental.M1 for details; Gaziano et al., 2016). A total of 222, 518 unrelated participants were included, of both European (EUR, n=193, 744), and African (AFR, n=28, 774) ancestries. Second, to study longitudinal brain changes, data from the Lifespan Changes in Brain and Cognition (LCBC) study, part of the Lifebrain project, was used. This dataset represents an adult lifespan sample comprising 1, 430 MRI scans from 420 healthy participants aged 30 to 89 years (248 females; mean age = 63.7 [SD = 14.4]). A subset of participants (n = 229) with both quality-controlled genetic data and longitudinal brain change estimates, derived from full adult lifespan models (Roe et al., 2024), was included. Each participant contributed between 2 and 7 timepoints inclusive (median=3), with follow-up durations ranging from 55 days to 11 years and 1 month. All participants provided written informed consent, and the study received approval from the relevant institutional and ethics review boards (Roe et al., 2024; see Supplemental.M2 for more details). The genetic studies included 639, 063 participants of EUR (n=554, 186) and AFR (n=84, 877) ancestry, comprising seven genome-wide association study cohorts (Supplemental.T1; Deak et al., 2022).

### Opioid use traits

Opioid use disorder was used in observational analyses as a severe phenotype for physiological dependence. In MVP, lifetime cases were identified using International Classification of Diseases (ICD) 9/10 diagnostic codes in the linked electronic health records (EHR) (Supplemental.T2; Deak et al., 2022). Unscreened individuals who had no documented diagnosis of OUD were included as controls in MVP (Deak et al., 2022).

For Lifebrain study, opioid use was an exclusion criterion for the enrolment, assessed through a detailed screening process. During enrolment, participants were screened for drug use and medication history. They were asked whether they had ever regularly used drugs other than alcohol, used medication for mental health disorders, or taken any medication on a regular basis. A positive response to either of the first two criteria led to automatic exclusion. For regular medication use, participants were excluded if the medication was known to affect the central nervous system (CNS), such as opioids, benzodiazepines, or antidepressants.

### Outcome measurements

In the MVP analysis, given the uncertainty regarding the effects of opioid use on specific dementia subtypes, we employed an inclusive primary outcome of all-cause dementia. Cases were identified through the presence of relevant ICD codes in the EHR (Supplemental.T3, Topiwala et al. 2024). Prevalent cases (those with dementia at enrolment) were excluded from the observational analyses to reduce the risk of reverse causation. Secondary outcomes included dementia subtypes (Alzheimer’s disease and vascular dementia), stroke subtypes and neuroimaging endophenotypes of dementia and addiction.

In the polygenic risk score analysis using data from the Lifebrain consortium, we examined longitudinal changes in key brain regions, including the cortex, hippocampus, amygdala, white matter, accumbens, caudate, pallidum and thalamus which are commonly affected by neurodegeneration (Ávila-Villanueva et al., 2022; Herlinger et al., 2021). This analysis was conducted in a sample of healthy European adults with no history of opioid use, allowing us to assess whether genetic risk for OUD is associated with accelerated atrophy in these regions, potentially indicating an increased vulnerability to dementia-related neurodegenerative processes.

### Covariates

Potential confounds and effect modifiers were identified based on prior research (Qeadan et al., 2023; Zoe et al., 2022). Data on sociodemographic factors (age, sex, education, income), lifestyle behaviours (smoking), and physical and psychiatric health (body mass index, history of head injury, post-traumatic stress disorder, and diabetes) were collected through self-reported enrolment and lifestyle questionnaires (see Supplemental.M1 for more details). Comorbid substance use disorders were identified by the presence of relevant ICD codes in the EHR.

### Genetic variants

Genetic associations with OUD were sought from the largest published genome-wide association study (GWAS) (Deak et al., 2022). The latter meta-analysed data from seven cohorts (Supplemental.T1) encompassing a total of 639, 063 participants. Three independent (r2<0.1) single-nucleotide polymorphisms (SNPs) were associated at genome-wide significance (p<5×10^-8^) with OUD in EUR and used as instrumental variables in the main MR: rs79704991 and rs1799971 both located in the *OPRM1* gene, and rs11372849 located in *FURIN* gene. rs17514846, located in *FURIN*, was used as a proxy (r^2^>0.8) when rs11372849 was unavailable in the outcome dataset (Supplemental.T4).

Selection of genetic variants for cis-MR was biologically-rather than statistically-driven and restricted to genes encoding well-described opioid targets (Lambert et al., 2023). Two of the three OUD genome-wide significant SNPs (rs1799971 and rs79704991), located within *OPRM1*, which encodes the μ-opioid receptor were identified (Gill et al., 2024). Two, albeit weaker, instruments within *OPRD1*, which encodes the δ-opioid receptor (rs529520 and rs2236861; p=0.001 and p=8×10^-6^) were also identified (Supplemental.T5). There were no available cis-SNPs for *OPRK1*.

Genetic associations with all-cause dementia were obtained from a GWAS using MVP, of 451, 317 EUR ancestry individuals (n=25, 473 cases) (Topiwala et al., 2024). Other outcome GWAS for secondary analyses included: clinically diagnosed Alzheimer’s disease (Kunkle et al., 2019, n=35, 274 cases), vascular dementia (n=8, 702 cases, Fongang et al., 2024), and in view of the limited number of cases in the vascular dementia GWAS, stroke as a relevant cerebrovascular phenotype (Mishra et al., 2022; n=73, 652 cases)(Supplemental.T6). Genetic associations with cross-sectional brain structure endophenotypes were identified in a UK Biobank GWAS (n=33, 224, Smith et al, 2021).

### Statistical analyses

#### Observational analysis

Cox proportional hazards models were employed to assess observational associations between OUD and incident dementia, adjusting for potential confounds, stratified by ancestry. Follow-up length was defined as the interval between enrolment (when covariates were measured) and either the first date of diagnosis, death or last data collection (December 2019) – whichever came first. Assumptions of proportionality of hazards were checked formally with time interactions and violations handled by stratifying on covariates of no interest.

#### Genetic analyses

Two-sample MR was used to estimate associations between genetically-predicted OUD liability and brain health outcomes. The inverse variance weighted (IVW) method served as the primary MR analysis, with additional robust methods employed to test the strength of causal inference. Analyses were conducted using *TwoSampleMR* (version 0.6.3) and *MendelianRandomization*. Variant harmonization ensured associations between SNPs and exposure/outcomes reflected the same allele. MR estimates were scaled to represent a doubling in prevalence of OUD, to aid interpretability (Burgess et al., 2018). Multiple testing correction was performed using a 10% false discovery rate (FDR) (Benjamini and Hochberg, 1995). Further analyses were performed to understand aetiological pathways, including: 1) cis-MR, restricting to genetic instruments to those within opioid receptor genes (*OPRM1* and *OPRD1*); 2) Bayesian colocalization to test for confounding by linkage disequilibrium using the *coloc* package (version 5.2.3, see Supplementary Figures) (Wallace et al., 2020); 3) polygenic risk score analyses to test whether genetic propensity to OUD might be linked to faster brain atrophy over time in individuals with no opioid exposure. Colocalization quantifies the likelihood of a hypothesis given the data, of a shared causal variant (PP.H4) between the exposure and outcome at the exposure gene locus (Zuber et al., 2022). A threshold of PP.H4 + PP.H3>0.5 and PP.H4 / (PP.H4 + PP.H3)>0.5 was considered evidence of colocalizing signals. Conversely, the posterior probability for distinct variants (PP.H3), refers to the likelihood that the genetic variants influencing an exposure and an outcome are distinct but in close proximity, often due to linkage disequilibrium, suggesting genetic confounding through variants in linkage disequilibrium. Other hypotheses tested included PP.H0, which represents the probability of no association with either trait, PP.H1, the probability of an association with the exposure only, and PP.H2, the probability of an association with the outcome only. Summary statistics from a large OUD GWAS were used to determine score weights for PRS (Deak et al). We computed PRS-OUD using both a stringent genome-wide significant and more relaxed p value threshold (pstringent<5×10^-8^ and prelaxed<0.1). Linear models were used, with brain structural change measures operationalized as the random individual-specific slopes estimated by fitting a nonlinear term of age using Generalized Additive Mixed Models (GAMMs) on adult lifespan data. These models were corrected for the individual’s mean age across timepoints, sex, number of timepoints, the first 10 genetic ancestry principal components, scanner type, and intracranial volume (Roe et al., 2024). Random slopes are then interpretable as the degree of brain change relative to the expected change given a person’s age, which has been shown to correlate with polygenic scores for OUD in these data.

## Results

### Observational analyses

Amongst 222, 518 MVP participants of EUR ancestry (n=193, 744; Supplemental.T7), 6, 905 individuals (3.6%) had a diagnosis of OUD. The prevalence of OUD was higher in the AFR group (n=2, 494; 8.7%; Supplemental.T8) compared to the EUR group. 7, 370 from the EUR group and 1, 027 from the AFR group developed incident all-cause dementia over a follow-up period of up to 9 years (mean 4.3 years). Individuals with OUD were younger (mean age = 57.9[SD=10.8]), more likely to be male (91.3%) and more likely to be daily smokers (40.6%). Individuals with OUD were also overrepresented in lower education and income categories compared to controls (Table 1). Alcohol (66.5%) and cannabis (30.2%) use disorders were more prevalent amongst those with OUD, especially in AFR individuals (Table 1).

**Table 1:**
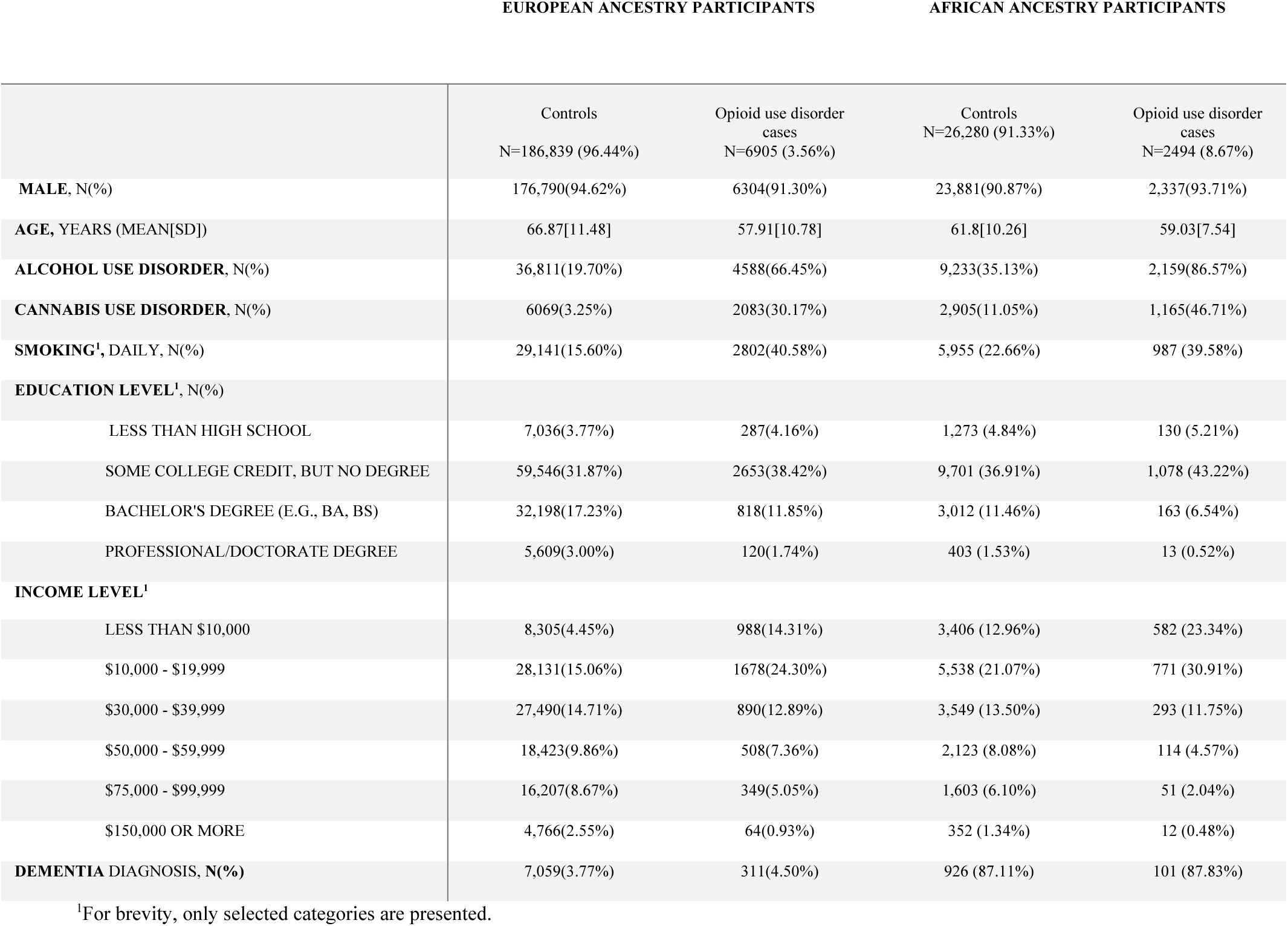
Demographic and clinical characteristics by opioid use disorder status in European(EUR) and African(AFR) ancestry individuals in Million Veteran Program (N = 222, 518). Complete data for European ancestry individuals are provided in Supplemental.T7-EUR, and for African ancestry individuals in Supplemental.T8-AFR.

In conventional observational analyses, participants with a diagnosis of OUD had a significantly higher incidence of all-cause dementia in both EUR (HR=1.56, 95% CI [1.39, 1.76], p=2.23×10^-13^) and AFR ancestry groups (HR=1.27, 95% CI [1.02, 1.59], p=0.03) (Fig. 1), after adjustment for the identified confounders. Significant positive associations with OUD were also observed in the EUR group for Alzheimer’s disease (HR = 1.40, 95% CI [1.04, 1.87], p = 0.02) and vascular dementia (HR = 1.49, 95% CI [1.19, 1.86], p = 0.0004), unlike in the lower powered AFR analyses (Fig. 1).

**Fig. 1:**
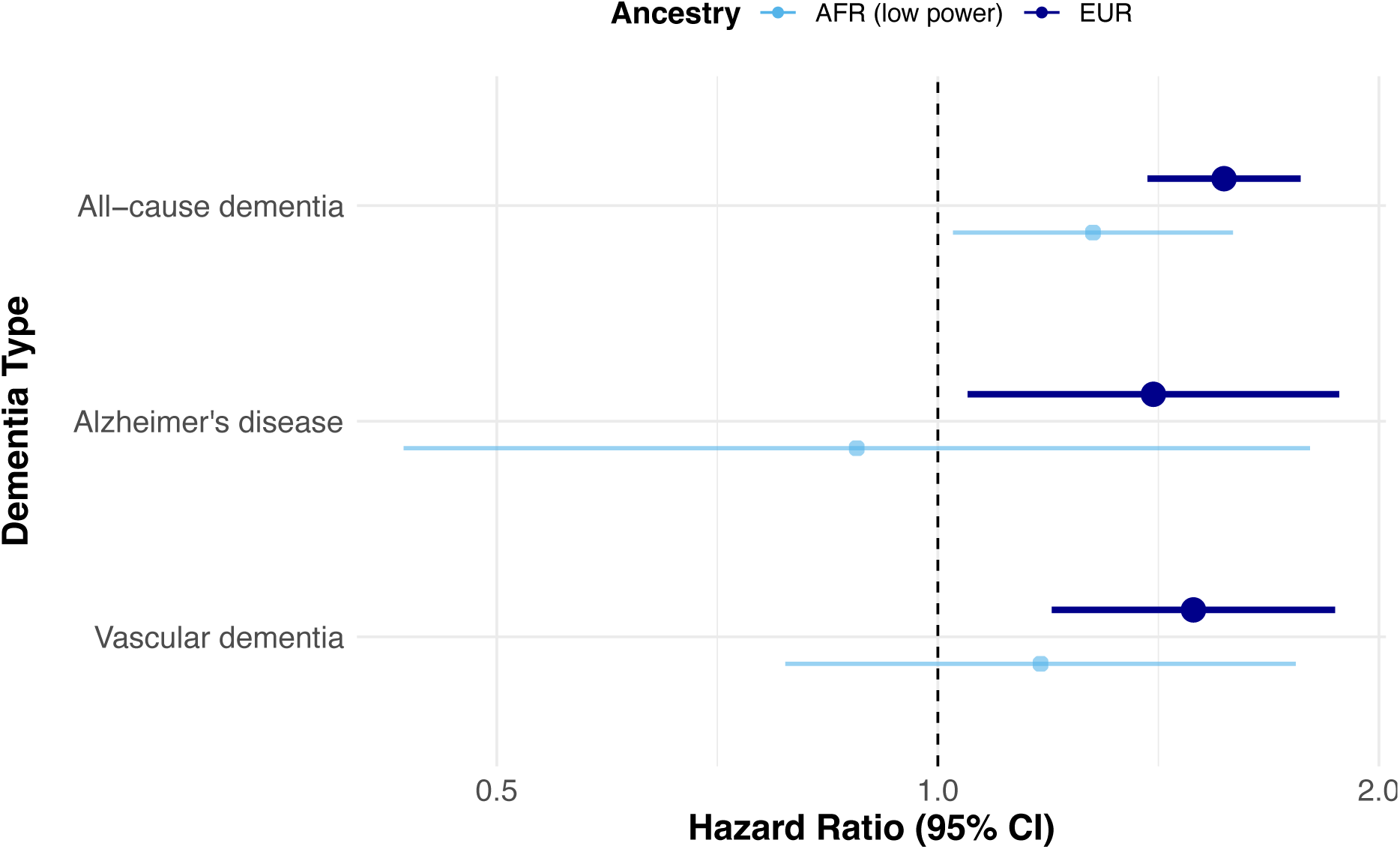
Observational associations between opioid use disorder and dementia subtypes in Million Veteran Program (n=222, 518), stratified by European (EUR) and African (AFR) ancestry. Dots represent hazard ratios and lines their 95% confidence intervals. Estimates were generated using Cox proportional hazards regression. Models were adjusted for: age, sex, educational qualification, income, smoking, alcohol and cannabis use disorder, depressive and post-traumatic stress disorder symptoms, body mass index.

### Genetic analyses

Three independent variants at genome-wide significance for OUD were used as instrumental variables in the primary MR analysis. Higher genetic propensity to OUD was significantly associated with all-cause dementia (IVW OR=1.77[1.43, 2.19], p=1.69×10⁻⁷) (Fig. 2).To interpret these findings, the observed odds ratio corresponds to a genetic association where a doubling in the genetic predisposition to OUD prevalence is associated with a 77% higher risk of all-cause dementia. Results from robust methods were broadly similar, albeit with wider confidence intervals (Supplemental.T9 and Supplemental Fig. 1A). In particular, genetic associations remained significant in multivariable MR, when controlling for genetically proxied chronic pain (MVMR IVW OR=1.82[1.18, 2.80], p<0.01), alcohol use disorder (MVMR IVW OR=1.81[1.41, 2.33], p<0.01), and cannabis use disorder (MVMR IVW OR=1.87[1.35, 2.61], p<0.01) (Supplemental.T10).

**Fig. 2:**
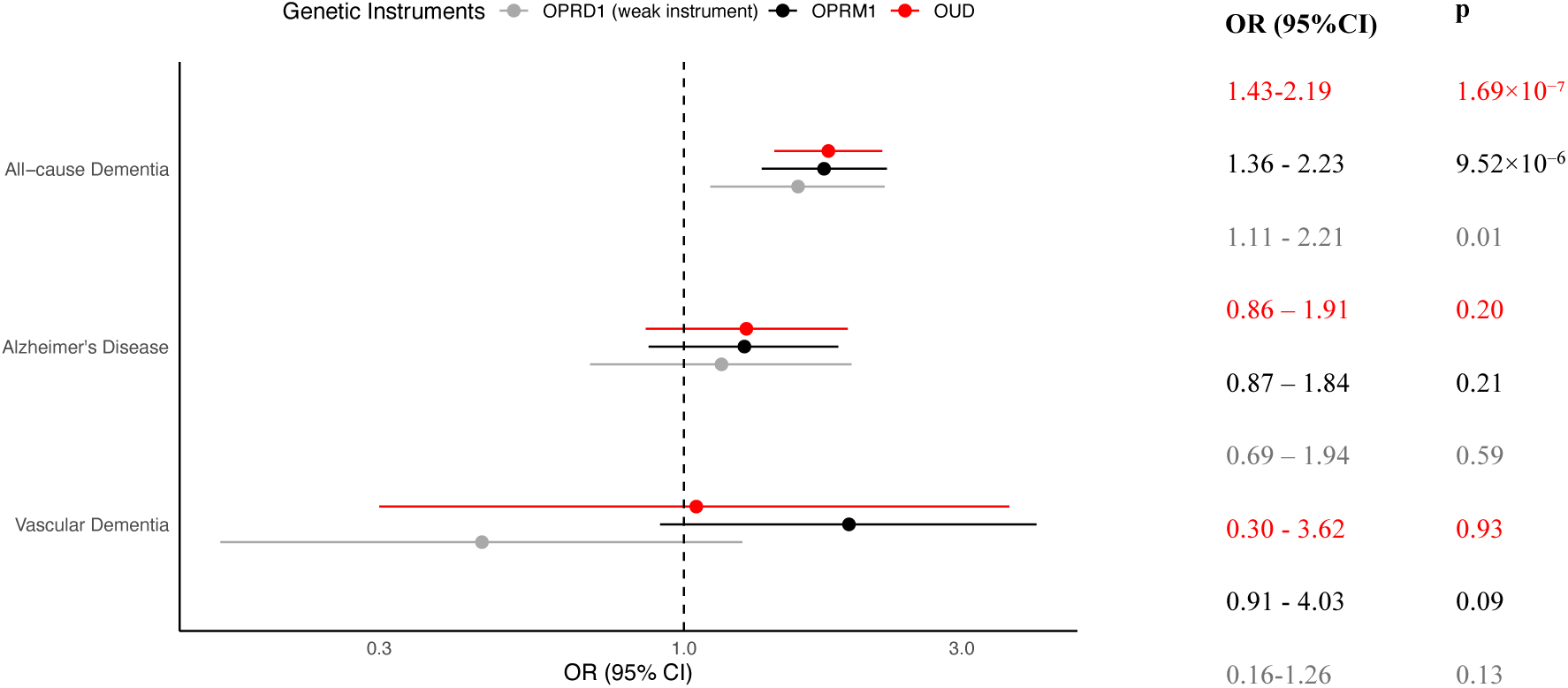
Genetic associations between opioid use disorder (OUD) and opioid receptor genes and dementia, scaled to reflect genetic variants associated with a doubling of OUD prevalence. Results are shown with 95% confidence intervals (95%CI) and p-values (p) in corresponding colors: OUD in red, OPRM1 in black, and OPRD1 in gray for all-cause dementia, Alzheimer’s disease, and vascular dementia separately. Estimates were generated using the inverse variance weighted method. Source genome-wide association studies used to derive genetic associations are described in Supplemental.T6.

In cis-MR analysis where we restricted genetic instruments to within opioid target genes, we identified two independent variants as genetic instruments within *OPRM1* (rs1799971 and rs79704991), and two variants as instruments within *OPRD1* (rs529520 and rs2236861) (Supplemental.T5). Both *OPRM1* (IVW OR=1.75[1.36, 2.23], p=9.5×10⁻⁶) and *OPRD1* (IVW OR=1.57[1.11, 2.21], p=0.01) instruments were associated with increased all-cause dementia risk (Fig. 2 & Supplemental Fig. 2 A and B). Colocalization analysis (Supplemental Fig. 3 A and B), indicated that a shared genetic association signal between *OPRM1* instruments (PP.H0=1.25×10^-4^, PP.H1=0.06, PP.H2=1.88×10^-5^, PP.H3=0.01, PP.H4=0.93) and dementia was the most likely scenario. In contrast, a low likelihood of a shared causal variant between *OPRD1* instruments (PP.H0=0.35, PP.H1=0.40, PP.H2=0.01, PP.H3=0.01, PP.H4=0.22) and dementia was found, which may be attributable to low statistical power (Supplemental Fig. 3 C and D).

We performed secondary MR analyses of genetically proxied OUD propensity and different dementia subtypes, as well as relevant endophenotypes of dementia. Estimates were directionally concordant for both Alzheimer’s disease (IVW OR=1.28[0.86, 1.91], p=0.20) and vascular dementia (IVW OR=1.05[0.30, 3.62], p=0.93), but with wide confidence intervals (Fig. 2). See Supplemental.T9 and Supplemental Fig. 1B and 1C for results using robust methods. Restricting genetic instruments to variants within *OPRM1* did not substantially alter associations (Supplemental Fig. 4 A and B; C and D for *OPRD1*). There was no significant association with any stroke phenotype using the genome-wide SNPs (ischemic stroke IVW OR = 1.07[0.70, 1.63], p=0.74; cardioembolic stroke IVW OR = 0.68[0.32, 1.45], p=0.31;large artery stroke IVW OR = 1.25[0.21, 7.31], p=0.8;small vessel stroke IVW OR= 1.50[0.77, 2.90], p=0.22) (Supplemental Fig. 6, Supplemental.T11-T12, Supplemental Fig. 5 A-D).

There were no significant associations with any cross-sectional neuroimaging dementia or addiction phenotypes examined (Supplemental Fig. 7., see Supplemental Fig. 8 for more information), including the left hippocampus (IVW beta= −0.22[-0.47, 0.02], p=0.07), left amygdala (IVW beta=0.05[-0.20, 0.31], p=0.68) and right ventral striatum(IVW beta=0.18[−0.07, 0.43], p=0.15) (Supplemental.T13 & T14). In individuals without opioid exposure, no associations were observed between genetic propensity to OUD, estimated using either a stringent or more relaxed OUD-PRS, and longitudinal brain atrophy. Regions tested included global cortical (PRSrelaxed *beta=0.05, t=0.8, p=0.4)* and subcortical regions including the hippocampus (PRSrelaxed *beta=-0.05, t=-0.7, p=0.4)* and amygdala (PRSrelaxed *beta= −0.1, t= −1.6, p=0.*09) (Supplemental.T15).

## Discussion

In this large-scale study, we triangulated research designs to investigate causal links between OUD and dementia, as well as the underlying biological mechanisms. Both observational and genetic analyses consistently supported a role for OUD in increasing dementia risk. Genetic evidence highlighted an aetiological role for perturbation of the μ-opioid receptor, which may mediate this relationship.

Our observational associations between OUD and dementia are consistent with some prior studies (Neelamegam et al., 2021; Levine et al., 2022; Sun et al., 2023; Pourhadi et al., 2024), but not all (Rapeli et al., 2006; Pask et al., 2019; Højsted et al., 2012). Differences could be accounted for by smaller sample sizes of earlier studies, which could have limited their statistical power to detect associations. Additionally, varying methods of OUD ascertainment, including self-report or clinical diagnosis, as well as differing dementia outcomes have been employed. Genetic propensity to OUD significantly associated with dementia in MR analyses, providing some evidence for a causal relationship. This was further strengthened by analyses that restricted genetic instruments to biologically relevant opioid receptor genes, as well as colocalization analyses, which suggested that these associations were not explained by genetic confounding but were likely driven by perturbations downstream of OUD. Notably, we found no evidence of genetic associations with brain structure in individuals with minimal or no opioid exposure, suggesting that the effects on dementia are driven by direct opioid exposure rather than shared genetic predisposition.

A key novelty of this study is the investigation of potential aetiological pathways through opioid receptor genes, *OPRM1* and *OPRD1*, which encode the μ- and δ-opioid receptors, respectively. While previous studies have explored the role of these receptors in the development of opioid dependence (Al-Hasani et al., 2013; Darcq et al., 2018; Dhaliwal et al., 2023), none have directly examined their individual connection to dementia. Associations were strongest with genetically-proxied μ-opioid receptor perturbation. The μ-opioid receptor is the primary modulator of analgesic effects of opioids, (Herman et al., 2024; Dhaliwal., 2024) and is also thought to modulate key neurobiological processes such as neurotransmitter release, synaptic plasticity, and neuroinflammation (Xu et al., 2023).

The mechanisms by which opioid use or dependence could result in dementia are currently unclear. Chronic opioid exposure can lead to a range of adverse outcomes, including atherosclerosis (Toska et al., 2023) and infection (Schwetz et al., 2019). Respiratory depression, hypoxia (Baldo et al., 2022;Kapil et al., 2018), and hypotension (Chen et al., 2015) could reduce blood flow to the brain and increase the likelihood of ischemic events. Although we did not find a significant positive association between OUD and stroke, previous studies suggest potential cerebrovascular consequences of opioid use (Janszky et al., 2021; Krantz et al., 2021), which may critically contribute to cognitive decline and dementia (Kalaria et al., 2016). Further investigation is needed to better understand these relationships. In spite of associations with dementia outcomes, we found no clear evidence for genetic associations with brain structure in individuals with minimal or no opioid exposure. This lack of association contrasts with previous smaller observational studies that have reported structural brain alterations in individuals with OUD or chronic opioid use (Schmidt et al., 2020; Upadhyay et al., 2010; Herlinger et al., 2021). Taken together, we hypothesize that effects are restricted to opioid users, and that genetic propensity to OUD does not relate to dementia (genetic confounding). Higher opioid exposure in MVP compared to UKB may explain why MR estimates of dementia outcomes were significant in contrast to brain structure.

A major strength of this study is the integration of large-scale observational data with genetic analyses. This triangulated approach reduces biases typically associated with observational studies, such as confounding and reverse causality, and strengthens causal inference, offering a more comprehensive understanding of the long-term effects of opioid use on brain health (Munafo 2018, Nature). The use of a large and diverse sample from the MVP, alongside genome-wide association studies, enhances the robustness and generalizability of our findings. Additionally, we examined a range of dementia outcomes, including all-cause dementia, Alzheimer’s disease, vascular dementia and relevant endophenotypes. We conducted further analyses to understand aetiological pathways, including mechanism-specific cis-MR, colocalization and employed PRS.

Several limitations should be noted. First, our study had greater power to detect associations in participants of EUR ancestry compared to AFR. Second, in observational analyses we were unable to investigate potentially important moderating factors, for example duration of OUD, specific opioid used, use of prescription or illicit opioids, or method of administration. MR depends on assumptions, and not all of these can be tested (Burgess et al., 2023). The MR estimates reflect the lifelong effects of genetically-proxied exposure, which cannot be equated to the effects of opioid use later in life (Burgess et al., 2017; Gill et al., 2021). Lastly, the variants mapped to *OPRD1* met only a lower threshold for suggestive statistical significance for association with OUD, in contrast to those for *OPRM1* (Kember et al., 2022; Deak et al., 2022; Zhou et al., 2020). This may explain why we found stronger links for the μ-opioid receptor and may not reflect actual differences in biology, but rather differences in the statistical power of the analysis.

## Conclusion

We found genetic evidence to support the hypothesis that OUD causally impacts dementia risk. The μ-opioid receptor (*OPRM1*) was implicated as a key biological pathway. These findings highlight the need for caution in prescribing opioids for long-term use and call for enhanced pharmacovigilance. Future research should explore the biological mechanisms linking *OPRM1* pathways to cognitive decline.

## Supporting information

Supplementary Figures: Additional visual data and analyses supporting the main findings of the study.

Supplementary Materials: Additional information supporting the main study.

Supplementary Tables: Additional data tables supporting the findings of the study.

## Data Availability

All data produced in the present study are available upon reasonable request to the authors

## Acknowledgments

K.P.E. is funded by the HDH Wills 1965 Charitable Trust (Registered Charity in England & Wales, No: 1117747). A.T. is supported by a Wellcome Trust fellowship (216462/Z/19/Z) https://wellcome.org/. S.B. is funded by the Wellcome Trust (grant number 225790/Z/22/Z) and UK Research and Innovation Medical Research Council (grant number MC_UU_00040/01). J.D.D. receives support from the National Institute on Drug Abuse (award K01DA058807). VK is supported by the Wellcome Trust (225790/Z/22/Z) and the United Kingdom Research and Innovation Medical Research Council (MC_UU_00040/01).D.G. is the Chief Executive Officer of Sequoia Genetics, a private limited company that collaborates with investors, pharmaceutical companies, biotech firms, and academic institutions to leverage genetic data for drug discovery and development. D.G. also holds interests in several biotechnology companies.

## References

Ahmad, M. A., Kareem, O., Khushtar, M., Akbar, M., Haque, M. R., Iqubal, A., Haider, M. F., Pottoo, F. H., Abdulla, F. S., Al-Haidar, M. B., & Alhajri, N. (2022). Neuroinflammation: A Potential Risk for Dementia. International Journal of Molecular Sciences, 23(2), 616. 10.3390/ijms23020616

Al-Hasani, R., & Bruchas, M. R. (2011). Molecular Mechanisms of Opioid Receptor-dependent Signaling and Behavior. Anesthesiology, 115(6), 1363–1381. 10.1097/ALN.0b013e318238bba6

Aquilani, R., Cotta Ramusino, M., Maestri, R., Iadarola, P., Boselli, M., Perini, G., Boschi, F., Dossena, M., Bellini, A., Buonocore, D., Doria, E., Costa, A., & Verri, M. (2023). Several dementia subtypes and mild cognitive impairment share brain reduction of neurotransmitter precursor amino acids, impaired energy metabolism, and lipid hyperoxidation. Frontiers in Aging Neuroscience, 15. 10.3389/fnagi.2023.1237469

Badr, M. Y., Gad, E. A. E., Mubarak, A. A. E., El-Heneedy, Y. A. A., Ibrahim, A. M., Belal, A. A. E., & Deep, F. A. el. (2023). Impact of tramadol and heroin abuse on electroencephalography structure and cognitive functions. Middle East Current Psychiatry, 30(1), 92. 10.1186/s43045-023-00365-7

Baldo, B. A., & Rose, M. A. (2022). Mechanisms of opioid-induced respiratory depression. Archives of Toxicology, 96(8), 2247–2260. 10.1007/s00204-022-03300-7

Bhatia, G., Ganesh, R., & Kulkarni, A. (2023). Cognitive impairment in opioid use disorders: Is there a case for use of nootropics? Psychiatry Research, 326, 115335. 10.1016/j.psychres.2023.115335

Bourgault, Z., Rubin-Kahana, D. S., Hassan, A. N., Sanches, M., & le Foll, B. (2022). Multiple Substance Use Disorders and Self-Reported Cognitive Function in U.S. Adults: Associations and Sex-Differences in a Nationally Representative Sample. Frontiers in Psychiatry, 12. 10.3389/fpsyt.2021.797578

Burgess, S., Davey Smith, G., Davies, N. M., Dudbridge, F., Gill, D., Glymour, M. M., Hartwig, F. P., Kutalik, Z., Holmes, M. v., Minelli, C., Morrison, J. v., Pan, W., Relton, C. L., & Theodoratou, E. (2023). Guidelines for performing Mendelian randomization investigations: update for summer 2023. Wellcome Open Research, 4, 186. 10.12688/wellcomeopenres.15555.3

Burgess, S., Small, D. S., & Thompson, S. G. (2017). A review of instrumental variable estimators for Mendelian randomization. Statistical Methods in Medical Research, 26(5), 2333–2355. 10.1177/0962280215597579

Butelman, E. R., Goldstein, R. Z., Nwaneshiudu, C. A., Girdhar, K., Roussos, P., Russo, S. J., & Alia-Klein, N. (2023). Neuroimmune Mechanisms of Opioid Use Disorder and Recovery: Translatability to Human Studies, and Future Research Directions. Neuroscience, 528, 102–116. 10.1016/j.neuroscience.2023.07.031

Centers for Disease Control and Prevention. (2022). Provisional drug overdose death counts. Retrieved from https://www.cdc.gov/nchs/nvss/vsrr/drug-overdose-data.htm

Chen, A., & Ashburn, M. A. (2015). Cardiac Effects of Opioid Therapy. Pain Medicine, 16(suppl 1), S27–S31. 10.1111/pme.12915

Compton, W. M., Valentino, R. J., & DuPont, R. L. (2021). Polysubstance use in the U.S. opioid crisis. Molecular Psychiatry, 26(1), 41–50. 10.1038/s41380-020-00949-3

Cornford, C. S., Mason, J. M., & Inns, F. (2011). Deep vein thromboses in users of opioid drugs: incidence, prevalence, and risk factors. The British Journal of General Practice: The Journal of the Royal College of General Practitioners, 61(593), e781–6. 10.3399/bjgp11X613115

Cuitavi, J., Andrés-Herrera, P., Meseguer, D., Campos-Jurado, Y., Lorente, J. D., Caruana, H., & Hipólito, L. (2023). Focal mu-opioid receptor activation promotes neuroinflammation and microglial activation in the mesocorticolimbic system: Alterations induced by inflammatory pain. Glia, 71(8), 1906–1920. 10.1002/glia.24374

Darcq, E., & Kieffer, B. L. (2018). Opioid receptors: drivers to addiction? Nature Reviews Neuroscience, 19(8), 499–514. 10.1038/s41583-018-0028-x

Deak, J. D., Zhou, H., Galimberti, M., Levey, D. F., Wendt, F. R., Sanchez-Roige, S., Hatoum, A. S., Johnson, E. C., Nunez, Y. Z., Demontis, D., Børglum, A. D., Rajagopal, V. M., Jennings, M. v., Kember, R. L., Justice, A. C., Edenberg, H. J., Agrawal, A., Polimanti, R., Kranzler, H. R., & Gelernter, J. (2022). Genome-wide association study in individuals of European and African ancestry and multi-trait analysis of opioid use disorder identifies 19 independent genome-wide significant risk loci. Molecular Psychiatry, 27(10), 3970–3979. 10.1038/s41380-022-01709-1

Dhaliwal, A., & Gupta, M. (2023). Physiology, opioid receptor. In StatPearls [Internet]. StatPearls Publishing. Retrieved from https://www.ncbi.nlm.nih.gov/books/NBK546642/

Dhingra, L., Ahmed, E., Shin, J., Scharaga, E., & Magun, M. (2015). Cognitive Effects and Sedation. Pain Medicine, 16(suppl 1), S37–S43. 10.1111/pme.12912

Dublin, S., Walker, R. L., Gray, S. L., Hubbard, R. A., Anderson, M. L., Yu, O., Crane, P. K., & Larson, E. B. (2015). Prescription Opioids and Risk of Dementia or Cognitive Decline: A Prospective Cohort Study. Journal of the American Geriatrics Society, 63(8), 1519–1526. 10.1111/jgs.13562

Elliott, L. T., Sharp, K., Alfaro-Almagro, F., Shi, S., Miller, K. L., Douaud, G., Marchini, J., & Smith, S. M. (2018). Genome-wide association studies of brain imaging phenotypes in UK Biobank. Nature, 562(7726), 210–216. 10.1038/s41586-018-0571-7

Gaziano, J. M., Concato, J., Brophy, M., Fiore, L., Pyarajan, S., Breeling, J., Whitbourne, S., Deen, J., Shannon, C., Humphries, D., Guarino, P., Aslan, M., Anderson, D., LaFleur, R., Hammond, T., Schaa, K., Moser, J., Huang, G., Muralidhar, S., … O’Leary, T. J. (2016). Million Veteran Program: A mega-biobank to study genetic influences on health and disease. Journal of Clinical Epidemiology, 70, 214–223. 10.1016/j.jclinepi.2015.09.016

Gill, D., Dib, M.-J., Cronjé, H. T., Karhunen, V., Woolf, B., Gagnon, E., Daghlas, I., Nyberg, M., Drakeman, D., & Burgess, S. (2024). Common pitfalls in drug target Mendelian randomization and how to avoid them. BMC Medicine, 22(1), 473. 10.1186/s12916-024-03700-9

Gill, D., Georgakis, M. K., Walker, V. M., Schmidt, A. F., Gkatzionis, A., Freitag, D. F., Finan, C., Hingorani, A. D., Howson, J. M. M., Burgess, S., Swerdlow, D. I., Davey Smith, G., Holmes, M. v, Dichgans, M., Scott, R. A., Zheng, J., Psaty, B. M., & Davies, N. M. (2021). Mendelian randomization for studying the effects of perturbing drug targets. Wellcome Open Research, 6, 16. 10.12688/wellcomeopenres.16544.2

Guo, X., Hou, C., Tang, P., & Li, R. (2023). Chronic Pain, Analgesics, and Cognitive Status: A Comprehensive Mendelian Randomization Study. Anesthesia & Analgesia. 10.1213/ANE.0000000000006514

Gupta, K., Prasad, A., Nagappa, M., Wong, J., Abrahamyan, L., & Chung, F. F. (2018). Risk factors for opioid-induced respiratory depression and failure to rescue. Current Opinion in Anaesthesiology, 31(1), 110–119. 10.1097/ACO.0000000000000541

Herlinger, K., & Lingford-Hughes, A. (2022). Opioid use disorder and the brain: a clinical perspective. Addiction, 117(2), 495–505. 10.1111/add.15636

Herman, T. F., Cascella, M., & Muzio, M. R. (2024). Mu receptors. In StatPearls [Internet]. StatPearls Publishing. Retrieved from https://www.ncbi.nlm.nih.gov/books/NBK551554/

Hoffman, K. A., Ponce Terashima, J., & McCarty, D. (2019). Opioid use disorder and treatment: challenges and opportunities. BMC Health Services Research, 19(1), 884. 10.1186/s12913-019-4751-4

Hojsted, J., Paula Kurita, G., Kendall, S., Lundorff, L., Andrucioli de Mattos Pimenta, C., & Sjogren, P. (2012). Non-Analgesic Effects of Opioids: The Cognitive Effects of Opioids in Chronic Pain of Malignant and Non-Malignant Origin. An Update. Current Pharmaceutical Design, 18(37), 6116–6122. 10.2174/138161212803582522

Howell, B. A., Abel, E. A., Park, D., Edmond, S. N., Leisch, L. J., & Becker, W. C. (2021). Validity of Incident Opioid Use Disorder (OUD) Diagnoses in Administrative Data: a Chart Verification Study. Journal of General Internal Medicine, 36(5), 1264– 1270. 10.1007/s11606-020-06339-3

Hser, Y.-I., Mooney, L. J., Saxon, A. J., Miotto, K., Bell, D. S., & Huang, D. (2017). Chronic pain among patients with opioid use disorder: Results from electronic health records data. Journal of Substance Abuse Treatment, 77, 26–30. 10.1016/j.jsat.2017.03.006

Ishrat, S., Levey, D. F., Gelernter, J., Ebmeier, K. P., & Topiwala, A. (n.d.). Title: Association between cannabis use and brain imaging phenotypes in UK Biobank: an observational and Mendelian randomization study. 10.1101/2023.08.12.23294013

Janszky, I., Vardaxis, I., Lindqvist, B. H., Horn, J. W., Brumpton, B. M., Strand, L. B., Bakken, I. J., Alsnes, I. V., Romundstad, P. R., Ljung, R., Mukamal, K. J., & Sen, A. (2021). Assessing short-term risk of ischemic stroke in relation to all prescribed medications. Scientific Reports, 11(1), 21673. 10.1038/s41598-021-01115-7

Kalaria, R. N., Akinyemi, R., & Ihara, M. (2016). Stroke injury, cognitive impairment and vascular dementia. Biochimica et Biophysica Acta (BBA) - Molecular Basis of Disease, 1862(5), 915–925. 10.1016/j.bbadis.2016.01.015

Kember, R. L., Vickers-Smith, R., Xu, H., Toikumo, S., Niarchou, M., Zhou, H., Hartwell, E. E., Crist, R. C., Rentsch, C. T., Million Veteran Program Davis, L. K., Justice, A. C., Sanchez-Roige, S., Kampman, K. M., Gelernter, J., & Kranzler, H. R. (2022). Cross-ancestry meta-analysis of opioid use disorder uncovers novel loci with predominant effects in brain regions associated with addiction. Nature Neuroscience, 25(10), 1279–1287. 10.1038/s41593-022-01160-z

Koller, D., Friligkou, E., Stiltner, B., Pathak, G. A., Løkhammer, S., Levey, D. F., Zhou, H., Hatoum, A. S., Deak, J. D., Kember, R. L., Treur, J. L., Kranzler, H. R., Johnson, E. C., Stein, M. B., Gelernter, J., & Polimanti, R. (2024). Pleiotropy and genetically inferred causality linking multisite chronic pain to substance use disorders. Molecular Psychiatry, 29(7), 2021–2030. 10.1038/s41380-024-02446-3

Krantz, M. J., Palmer, R. B., & Haigney, M. C. P. (2021). Cardiovascular Complications of Opioid Use. Journal of the American College of Cardiology, 77(2), 205–223. 10.1016/j.jacc.2020.11.002

Kunkle, B. W., Grenier-Boley, B., Sims, R., Bis, J. C., Damotte, V., Naj, A. C., Boland, A., Vronskaya, M., van der Lee, S. J., Amlie-Wolf, A., Bellenguez, C., Frizatti, A., Chouraki, V., Martin, E. R., Sleegers, K., Badarinarayan, N., Jakobsdottir, J., Hamilton-Nelson, K. L., Moreno-Grau, S., … Pericak-Vance, M. A. (2019). Genetic meta-analysis of diagnosed Alzheimer’s disease identifies new risk loci and implicates Aβ, tau, immunity and lipid processing. Nature Genetics, 51(3), 414–430. 10.1038/s41588-019-0358-2

Lambert, D. G. (2023). Opioids and opioid receptors; understanding pharmacological mechanisms as a key to therapeutic advances and mitigation of the misuse crisis. BJA Open, 6, 100141. 10.1016/j.bjao.2023.100141

Ledger, J., Tetrault, J. M., & D’Onofrio, G. M. M. (2020). Opioid use disorder. Yale Medicine. Retrieved from https://www.yalemedicine.org/conditions/opioid-use-disorder

Levine, S. Z., Rotstein, A., Goldberg, Y., Reichenberg, A., & Kodesh, A. (2023). Opioid Exposure and the Risk of Dementia: A National Cohort Study. The American Journal of Geriatric Psychiatry, 31(5), 315–323. 10.1016/j.jagp.2022.05.013

Mega Vascular Cognitive Impairment and Dementia (MEGAVCID) consortium. (2024). A genome-wide association meta-analysis of all-cause and vascular dementia. Alzheimer’s & Dementia: The Journal of the Alzheimer’s Association, 20(9), 5973– 5995. 10.1002/alz.14115

Mishra, A., Malik, R., Hachiya, T., Jürgenson, T., Namba, S., Posner, D. C., Kamanu, F. K., Koido, M., le Grand, Q., Shi, M., He, Y., Georgakis, M. K., Caro, I., Krebs, K., Liaw, Y.-C., Vaura, F. C., Lin, K., Winsvold, B. S., Srinivasasainagendra, V., … Debette, S. (2022). Stroke genetics informs drug discovery and risk prediction across ancestries. Nature, 611(7934), 115–123. 10.1038/s41586-022-05165-3

Neelamegam, M., Zgibor, J., Chen, H., O’rourke, K., Bakour, C., Rajaram, L., & Anstey, K. J. (2021). The effect of opioids on the cognitive function of older adults: results from the Personality and Total Health through life study. Age and Ageing, 50(5), 1699–1708. 10.1093/ageing/afab048

Parikh, N. S., Merkler, A. E., & Iadecola, C. (2020). Inflammation, Autoimmunity, Infection, and Stroke. Stroke, 51(3), 711–718. 10.1161/STROKEAHA.119.024157

Pask, S., Dell’Olio, M., Murtagh, F. E. M., & Boland, J. W. (2020). The Effects of Opioids on Cognition in Older Adults With Cancer and Chronic Noncancer Pain: A Systematic Review. Journal of Pain and Symptom Management, 59(4), 871–893.e1. 10.1016/j.jpainsymman.2019.10.022

Pourhadi, N., Janbek, J., Gasse, C., Laursen, T. M., Waldemar, G., & Jensen-Dahm, C. (2024). Opioids and Dementia in the Danish Population. JAMA Network Open, 7(11), e2445904. 10.1001/jamanetworkopen.2024.45904

Qeadan, F., McCunn, A., Tingey, B., Price, R., Bobay, K. L., English, K., & Madden, E. F. (2023). Exploring the Association Between Opioid Use Disorder and Alzheimer’s Disease and Dementia Among a National Sample of the U.S. Population. Journal of Alzheimer’s Disease, 96(1), 229–244. 10.3233/JAD-230714

Ranapurwala, S. I., Alam, I. Z., Pence, B. W., Carey, T. S., Christensen, S., Clark, M., Chelminski, P. R., Wu, L., Greenblatt, L. H., Korte, J. E., Wolfson, M., Douglas, H. E., Bowlby, L. A., Capata, M., & Marshall, S. W. (2023). Development and validation of an electronic health records-based opioid use disorder algorithm by expert clinical adjudication among patients with prescribed opioids. Pharmacoepidemiology and Drug Safety, 32(5), 577–585. 10.1002/pds.5591

Rapeli, P., Kivisaari, R., Autti, T., Kähkönen, S., Puuskari, V., Jokela, O., & Kalska, H. (2006). Cognitive function during early abstinence from opioid dependence: a comparison to age, gender, and verbal intelligence matched controls. BMC Psychiatry, 6(1), 9. 10.1186/1471-244X-6-9

Rejdak, K., Sienkiewicz-Jarosz, H., Bienkowski, P., & Alvarez, A. (2023). Modulation of neurotrophic factors in the treatment of dementia, stroke and TBI: Effects of Cerebrolysin. Medicinal Research Reviews, 43(5), 1668–1700. 10.1002/med.21960

Roe, J. M., Vidal-Piñeiro, D., Sørensen, Ø., Grydeland, H., Leonardsen, E. H., Iakunchykova, O., Pan, M., Mowinckel, A., Strømstad, M., Nawijn, L., Milaneschi, Y., Andersson, M., Pudas, S., Bråthen, A. C. S., Kransberg, J., Falch, E. S., Øverbye, K., Kievit, R. A., Ebmeier, K. P., … Wang, Y. (2023). Accelerated brain change in healthy adults is associated with genetic risk for Alzheimer’s disease and uncovers adult lifespan memory decline. 10.1101/2023.10.09.559446

Saunders, G. R. B., Wang, X., Chen, F., Jang, S.-K., Liu, M., Wang, C., Gao, S., Jiang, Y., Khunsriraksakul, C., Otto, J. M., Addison, C., Akiyama, M., Albert, C. M., Aliev, F., Alonso, A., Arnett, D. K., Ashley-Koch, A. E., Ashrani, A. A., Barnes, K. C., … Vrieze, S. (2022). Genetic diversity fuels gene discovery for tobacco and alcohol use. Nature, 612(7941), 720–724. 10.1038/s41586-022-05477-4

Scherrer, J. F., Sullivan, M. D., LaRochelle, M. R., & Grucza, R. (2023). Validating opioid use disorder diagnoses in administrative data: a commentary on existing evidence and future directions. Addiction Science & Clinical Practice, 18(1), 49. 10.1186/s13722-023-00405-x

Schmidt, A., Vogel, M., Baumgartner, S., Wiesbeck, G. A., Lang, U., Borgwardt, S., & Walter, M. (2021). Brain volume changes after long-term injectable opioid treatment: A longitudinal voxel-based morphometry study. Addiction Biology, 26(4). 10.1111/adb.12970

Schwetz, T. A., Calder, T., Rosenthal, E., Kattakuzhy, S., & Fauci, A. S. (2019). Opioids and Infectious Diseases: A Converging Public Health Crisis. The Journal of Infectious Diseases, 220(3), 346–349. 10.1093/infdis/jiz133

Scuteri, D., Mantovani, E., Tamburin, S., Sandrini, G., Corasaniti, M. T., Bagetta, G., & Tonin, P. (2020). Opioids in Post-stroke Pain: A Systematic Review and Meta-Analysis. Frontiers in Pharmacology, 11. 10.3389/fphar.2020.587050

Smith, S. M., Douaud, G., Chen, W., Hanayik, T., Alfaro-Almagro, F., Sharp, K., & Elliott, L. T. (2021). An expanded set of genome-wide association studies of brain imaging phenotypes in UK Biobank. Nature Neuroscience, 24(5), 737–745. 10.1038/s41593-021-00826-4

Sulley, S., & Ndanga, M. (2020). Inpatient Opioid Use Disorder and Social Determinants of Health: A Nationwide Analysis of the National Inpatient Sample (2012-2014 and 2016-2017). Cureus, 12(11), e11311. 10.7759/cureus.11311

Sun, M., Chen, W.-M., Wu, S.-Y., & Zhang, J. (2023). Long-Term Opioid Use and Dementia Risk in Patients With Chronic Pain. Journal of the American Medical Directors Association, 24(9), 1420–1426.e2. 10.1016/j.jamda.2023.06.035

Tian, J., Jones, G., Lin, X., Zhou, Y., King, A., Vickers, J., & Pan, F. (2023). Association between chronic pain and risk of incident dementia: findings from a prospective cohort. BMC Medicine, 21(1), 169. 10.1186/s12916-023-02875-x

Tolomeo, S., Gray, S., Matthews, K., Steele, J. D., & Baldacchino, A. (2016). Multifaceted impairments in impulsivity and brain structural abnormalities in opioid dependence and abstinence. Psychological Medicine, 46(13), 2841–2853. 10.1017/S0033291716001513

Topiwala, A., Senior, - BMBCh, Levey - Assistant Professor, D. F., Zhou - Assistant Professor, H., Deak -PhD Postdoctoral Researcher, J. D., Ebmeier -MD Professor of Old, K. P., Bell - Senior Bioinformatician, S., Burgess - Principal Research Associate, S., Nichols -PhD Professor of Neuroimaging Statistics, T. E., Stein, M. B., & Gelernter - Professor, J. (n.d.). Alcohol use and risk of dementia in diverse populations. 10.1101/2024.05.20.24307606

Toska, E., & Mayrovitz, H. N. (2023). Opioid Impacts on Cardiovascular Health. Cureus. 10.7759/cureus.46224

Upadhyay, J., Maleki, N., Potter, J., Elman, I., Rudrauf, D., Knudsen, J., Wallin, D., Pendse, G., McDonald, L., Griffin, M., Anderson, J., Nutile, L., Renshaw, P., Weiss, R., Becerra, L., & Borsook, D. (2010a). Alterations in brain structure and functional connectivity in prescription opioid-dependent patients. Brain, 133(7), 2098–2114. 10.1093/brain/awq138

Upadhyay, J., Maleki, N., Potter, J., Elman, I., Rudrauf, D., Knudsen, J., Wallin, D., Pendse, G., McDonald, L., Griffin, M., Anderson, J., Nutile, L., Renshaw, P., Weiss, R., Becerra, L., & Borsook, D. (2010b). Alterations in brain structure and functional connectivity in prescription opioid-dependent patients. Brain, 133(7), 2098–2114. 10.1093/brain/awq138

Upp, L. A., & Waljee, J. F. (2020). The Opioid Epidemic. Clinics in Plastic Surgery, 47(2), 181–190. 10.1016/j.cps.2019.12.005

Verdejo-Garcia, A., Garcia-Fernandez, G., & Dom, G. (2019). Cognition and addiction . Dialogues in Clinical Neuroscience, 21(3), 281–290. 10.31887/DCNS.2019.21.3/gdom

Wallace, C. (2020). Eliciting priors and relaxing the single causal variant assumption in colocalisation analyses. PLOS Genetics, 16(4), e1008720. 10.1371/journal.pgen.1008720

Werner, C., & Kochs, E. (1996). Opioids and intracranial pressure. Current Opinion in Anaesthesiology, 9(5), 385–388.

Xu, Y., Chen, R., Zhi, F., Sheng, S., Khiati, L., Yang, Y., Peng, Y., & Xia, Y. (2023). δ-opioid Receptor, Microglia and Neuroinflammation. Aging and Disease, 14(3), 778. 10.14336/AD.2022.0912

Xue, X., Zong, W., Glausier, J. R., Kim, S.-M., Shelton, M. A., Phan, B. N., Srinivasan, C., Pfenning, A. R., Tseng, G. C., Lewis, D. A., Seney, M. L., & Logan, R. W. (2022). Molecular rhythm alterations in prefrontal cortex and nucleus accumbens associated with opioid use disorder. Translational Psychiatry, 12(1), 123. 10.1038/s41398-022-01894-1

Zhou, H., Rentsch, C. T., Cheng, Z., Kember, R. L., Nunez, Y. Z., Sherva, R. M., Tate, J. P., Dao, C., Xu, K., Polimanti, R., Farrer, L. A., Justice, A. C., Kranzler, H. R., Gelernter, J., & Veterans Affairs Million Veteran Program. (2020). Association of OPRM1 Functional Coding Variant With Opioid Use Disorder: A Genome-Wide Association Study. JAMA Psychiatry, 77(10), 1072–1080. 10.1001/jamapsychiatry.2020.1206

Zuber, V., Grinberg, N. F., Gill, D., Manipur, I., Slob, E. A. W., Patel, A., Wallace, C., & Burgess, S. (2022). Combining evidence from Mendelian randomization and colocalization: Review and comparison of approaches. The American Journal of Human Genetics, 109(5), 767–782. 10.1016/j.ajhg.2022.04.001

