## Supplementary Figures: Additional visual data and analyses supporting the main findings of the study. for "Opioid use disorder and brain health: observational and genetic associations"

**SUPPLEMENTARY FIGURES** 2


**Supplemental Fig. 1** 2
A- Scatterplot and Forest Plot of Genetic Associations Between OUD and All-cause dementia in MR Analysis 2
B- Scatterplot and Forest Plot of Genetic Association Between OUD and Alzheimer's disease in MR Analysis 2
C- Scatterplot and Forest Plot of Genetic Association Between OUD and vascular dementia in MR Analysis 3

**Supplemental Fig. 2** 3
A- Scatterplot and Forest Plot of Genetically-Proxied μ-Opioid Receptor Perturbation and Increased Risk of All-Cause Dementia 3
B- Scatterplot and Forest Plot of Genetically-Proxied δ - Opioid Receptor Perturbation and Increased Risk of All-Cause Dementia 4

**Supplemental Fig. 3** 5
A- Manhattan Plots Highlighting Association Signals for *OPRM1* and All-Cause Dementia on Chromosome 6 5
B- Colocalization Analysis of *OPRM1* and All-Cause Dementia: Association Signals, Prior Assumptions, and Posterior Probabilities Across Hypotheses 6
C- Manhattan Plots Highlighting Association Signals for *OPRD1* and All-Cause Dementia on Chromosome 1 7
D- Colocalization Analysis of *OPRD1* and All-Cause Dementia: Association Signals, Prior Assumptions, and Posterior Probabilities Across Hypotheses 8

**Supplemental Fig. 4** 9
A- Scatterplot and Forest Plot of Genetically-Proxied μ-Opioid Receptor Perturbation and Alzheimer's disease risk 9
B- Scatterplot and Forest Plot of Genetically-Proxied μ-Opioid Receptor Perturbation and Vascular dementia risk. 9
C- Scatterplot and Forest Plot of Genetically-Proxied δ -Opioid Receptor Perturbation and Alzheimer's disease risk. 10
D- Scatterplot and Forest Plot of Genetically-Proxied δ -Opioid Receptor Perturbation and Vascular dementia. 10

**Supplemental Fig. 5** 11
A- Scatterplot and Forest Plot of Genetic Association Between OUD and Ischemic Stroke in MR Analysis. 11
B- Scatterplot and Forest Plot of Genetic Association Between OUD and Large Artery Stroke in MR Analysis 11
C- Scatterplot and Forest Plot of Genetic Association Between OUD and Small Vessel Stroke in MR Analysis 12
D- Scatterplot and Forest Plot of Genetic Association Between OUD and Cardioembolic Stroke in MR Analysis 12

**Supplemental Fig. 6** 13
Genetic Association between OUD and Stroke Subtypes. 13

**Supplemental Fig. 7** 13
Genetic Association of OUD with Brain Structure Metrics in MR Analysis 13

**Supplemental Fig. 8** 14
Mendelian Randomization Analysis of Opioid Use Disorder and Brain Structure: Forest and Scatter Plots of Genetic Associations 14

### Supplementary figures

#### Supplemental Fig. 1

##### A- Scatterplot and Forest Plot of Genetic Associations Between OUD and All-cause dementia in MR Analysis

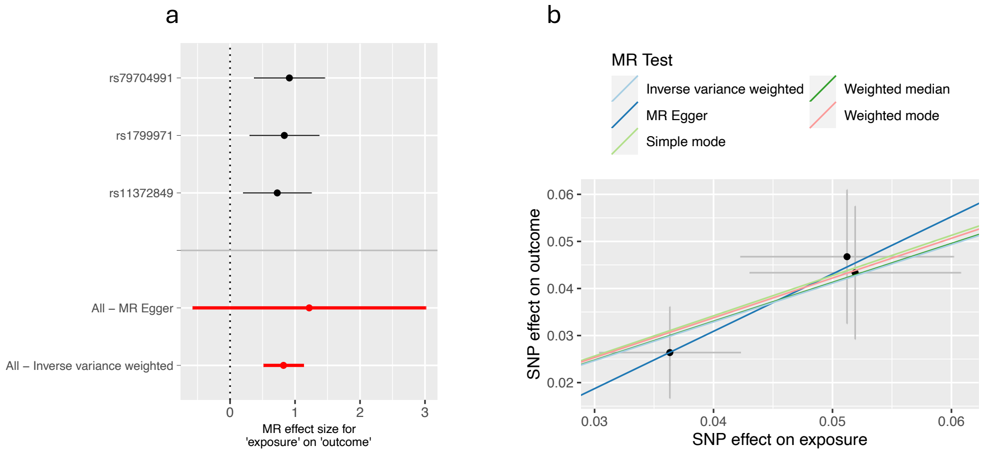


**MR analysis of the association between OUD and all-cause dementia. a** Forest plot illustrating the MR effect sizes of OUD on dementia, using Genome-wide significant (GWS) variants associated with OUD and MR methods. Each black point represents the estimated MR effect size for the association between OUD and dementia for individual SNPs (rs79704991, rs1799971, rs11372849), with black lines indicating their 95% confidence intervals. The red points and lines represent the overall MR estimates using the MR Egger and inverse variance weighted (IVW) methods, with the corresponding confidence intervals. A dotted line at zero indicates the null hypothesis of no effect. **b** MR scatter plot showing the association between OUD and all-cause dementia, with results from different MR methods. Each black point represents a SNP (Single Nucleotide Polymorphism), and the gray bars indicate the standard error of the mean (SEM). The dementia β value is the natural log of the odds ratio, where lower values correspond to a reduced likelihood of developing dementia. The OUD β is reported as normalized effect sizes (NESs), with larger values indicating a higher risk of OUD.

##### B- Scatterplot and Forest Plot of Genetic Association Between OUD and Alzheimer's disease in MR Analysis

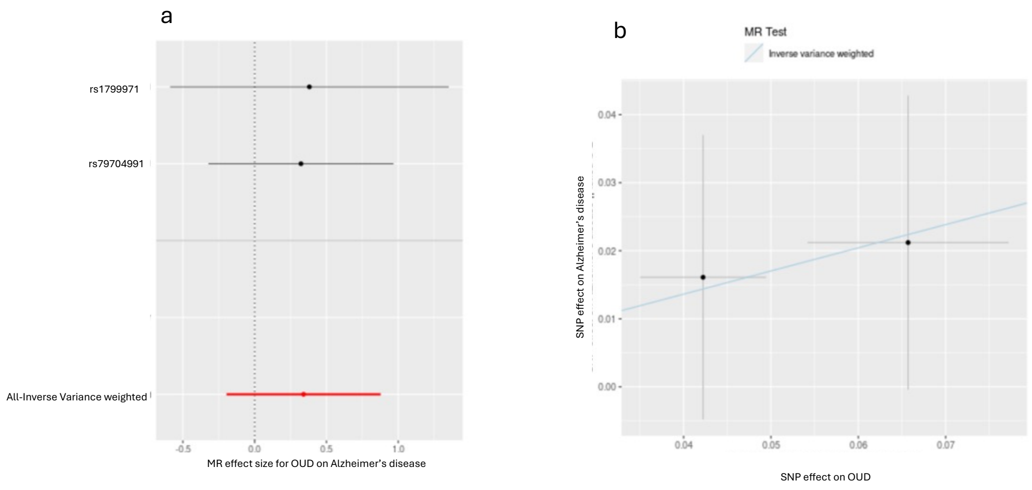


**MR Analysis of the Genetic Association Between Opioid Use Disorder (OUD) and Alzheimer's Disease. a** Forest plot illustrating the Mendelian randomization (MR) estimates for the association between OUD and Alzheimer's disease using the inverse variance weighted (IVW) method. The x-axis represents the MR effect size, with 95% confidence intervals shown for each estimate. **b** Scatterplot showing the SNP effects on OUD (x-axis) and Alzheimer's disease (y-axis). The regression line represents the IVW MR estimate, providing a visualization of the direction and strength of the association.

#### C- Scatterplot and Forest Plot of Genetic Association Between OUD and vascular dementia in MR Analysis

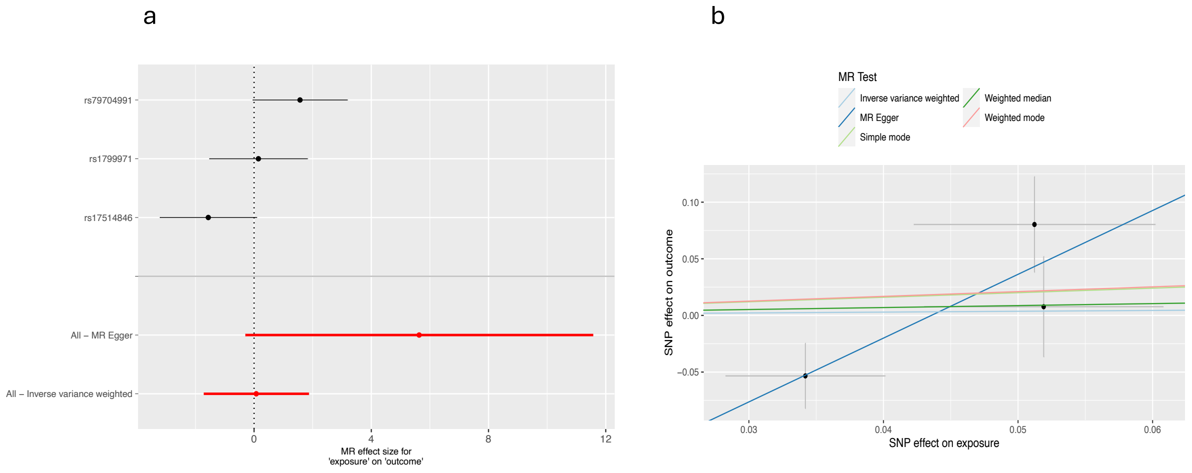


**MR Analysis of the Genetic Association Between Opioid Use Disorder (OUD) and Vascular dementia. a** Forest plot illustrating the Mendelian randomization (MR) estimates for the association between OUD and Vascular dementia using the MR-Egger and inverse variance weighted (IVW) methods. The x-axis represents the MR effect size, with 95% confidence intervals shown for each estimate. **b** Scatterplot showing the SNP effects on OUD (x-axis) and Vascular dementia (y-axis). The regression line represents the IVW MR estimate, illustrating the direction and strength of the genetic association.

### Supplemental Fig. 2

#### A- Scatterplot and Forest Plot of Genetically-Proxied μ-Opioid Receptor Perturbation and Increased Risk of All-Cause Dementia

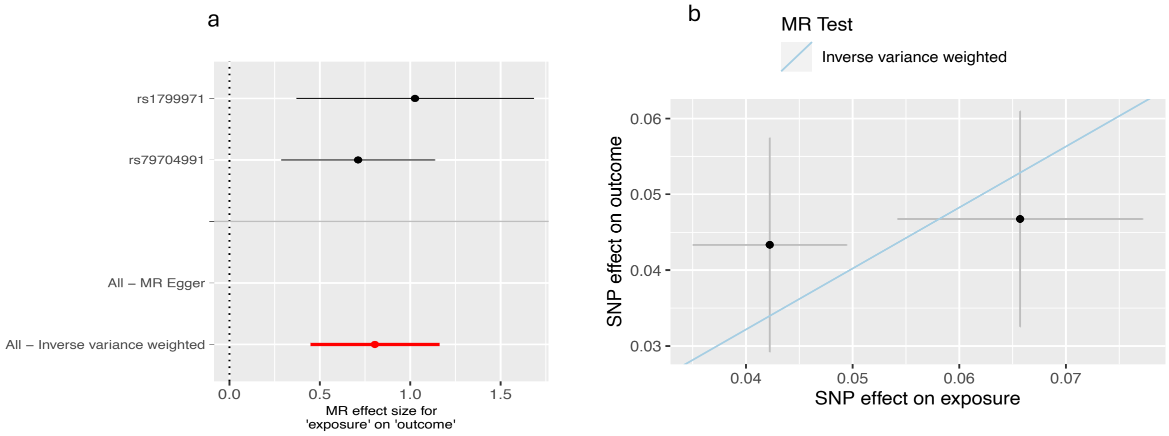


Cis-Mendelian randomization (MR) analysis of genetically-proxied μ-opioid receptor perturbation and increased all-cause dementia risk. Cis-acting SNPs within the *OPRM1* gene were selected, and genetic associations with opioid OUD were obtained from the largest published genome-wide association study (GWAS) (Deak et al., 2022). Genetic associations with all-cause dementia were derived from Topiwala et al., 2024. (a) Scatter plot showing cis-acting SNP-specific effects on OUD versus all-cause dementia risk. The slope of the regression line represents the causal effect estimated using the inverse variance weighted (IVW) method. (b) Forest plot displaying individual SNP-specific causal estimates and the overall causal estimates derived from MR-Egger and IVW methods. The red line indicates the overall effect size estimated using the IVW method, while the dashed vertical line denotes the null hypothesis (no causal effect). The results provide evidence for a causal relationship between μ-opioid receptor perturbation and all-cause dementia risk (IVW OR = 1.75 [1.36, 2.23]; $p = 9.5 \times 10^{-6}$).

#### B- Scatterplot and Forest Plot of Genetically-Proxied δ - Opioid Receptor Perturbation and Increased Risk of All-Cause Dementia

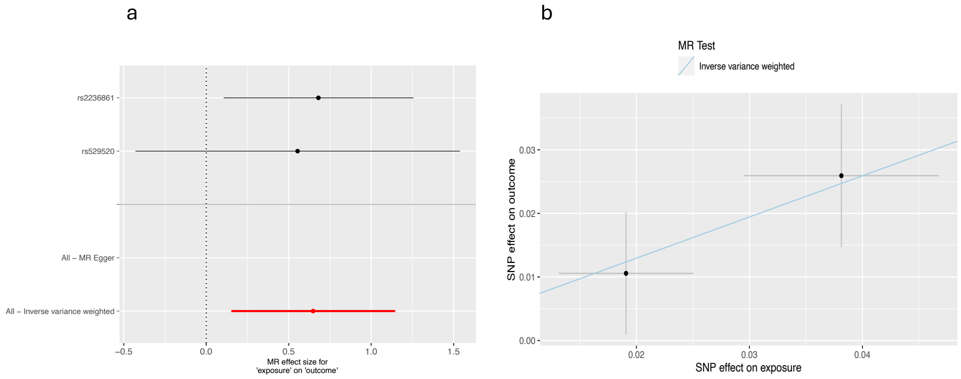


Cis-Mendelian randomization (MR) analysis of genetically-proxied δ-opioid receptor perturbation and increased all-cause dementia risk. Cis-acting SNPs within the *OPRD1* gene were selected, with genetic associations for OUD obtained from the largest published genome-wide association study (GWAS) (Deak et al., 2022) and genetic associations for all-cause dementia derived from Topiwala et al., 2024. (a) Scatter plot showing the effects of cis-acting SNPs on OUD (δ-opioid receptor perturbation) versus all-cause dementia risk. The slope of the regression line indicates the causal effect estimated using the inverse variance weighted (IVW) method. (b) Forest plot displaying individual SNP-specific causal estimates and the overall causal estimates obtained using MR-Egger and IVW methods. The red line represents the overall effect size estimated using the IVW method, while the dashed vertical line denotes the null hypothesis (no causal effect). The results provide evidence for a causal relationship between δ-opioid receptor perturbation and all-cause dementia risk (IVW OR = 1.57 [1.11, 2.21]; p = 0.01).

#### Supplemental Fig. 3

#### A- Manhattan Plots Highlighting Association Signals for *OPRM1* and All-Cause Dementia on Chromosome 6

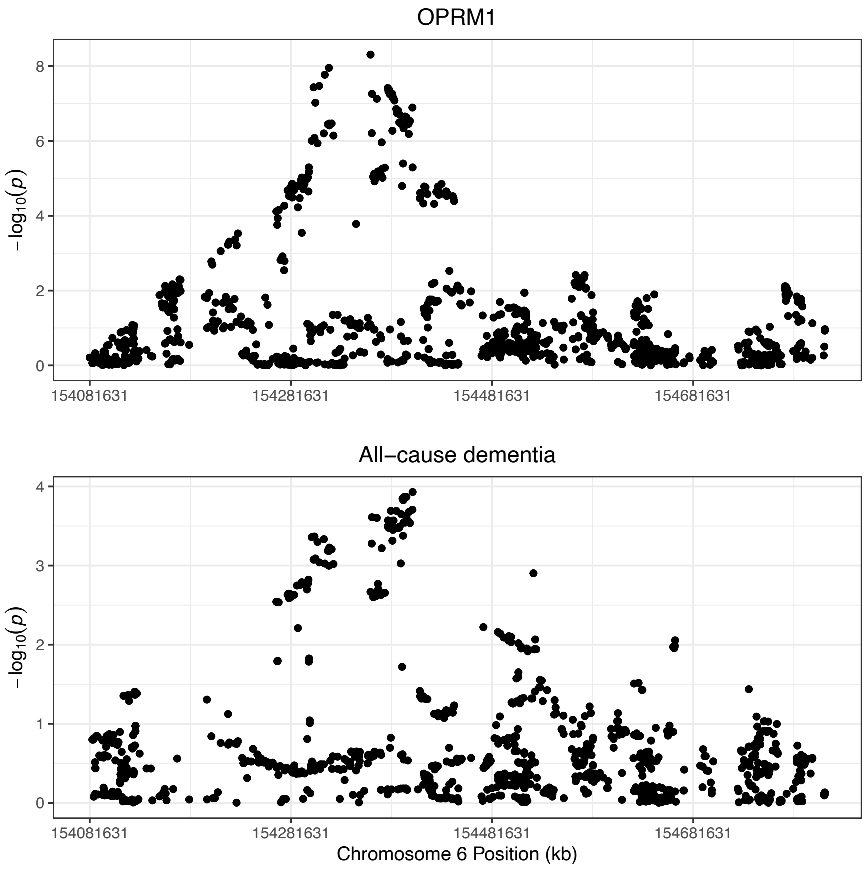


Manhattan plots showing association signals for *OPRM1* (top) and all-cause dementia (bottom) across Chromosome 6. Each point represents a genetic variant, with the y-axis indicating the -log10(p-value) of the association, and the x-axis representing genomic positions in kilobases (kb). Peaks in the plots highlight loci with strong associations, suggesting potential regions of interest for further investigation into shared or distinct genetic influences on these traits.

#### B- Colocalization Analysis of *OPRM1* and All-Cause Dementia: Association Signals, Prior Assumptions, and Posterior Probabilities Across Hypotheses

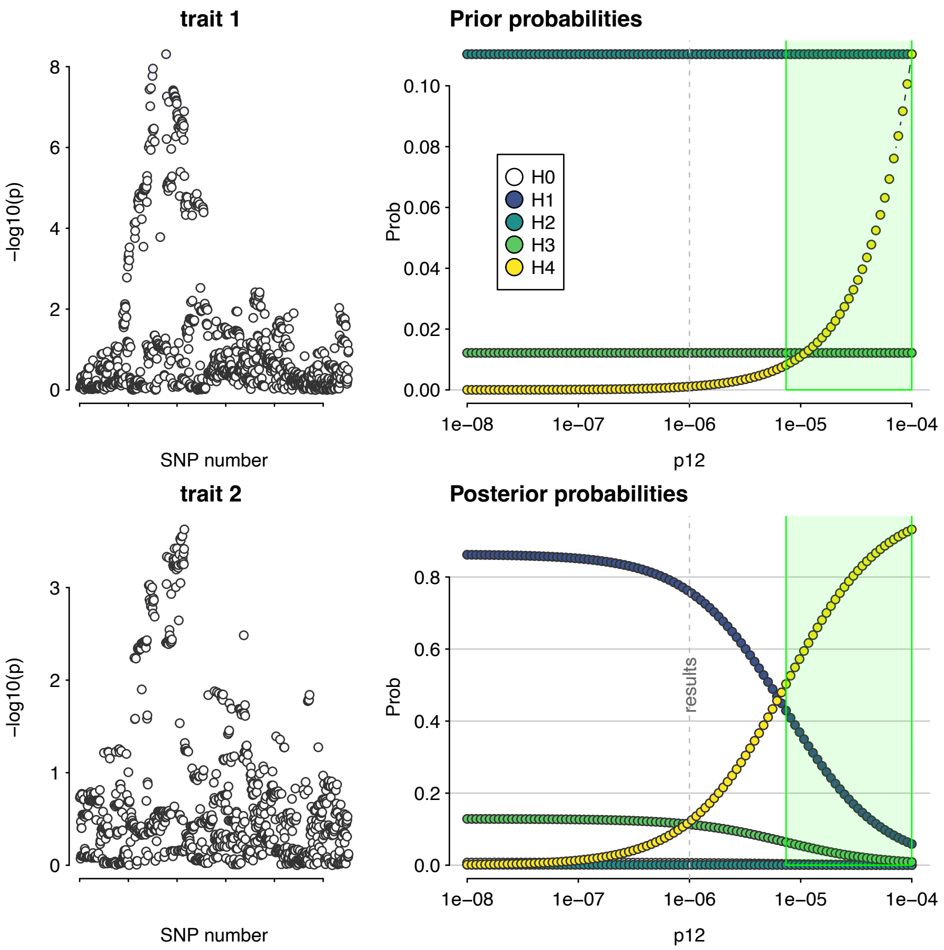


Colocalization analysis of *OPRM1* and all-cause dementia on Chromosome 6. Left panels: Manhattan plots showing the association signals (-log10(p-values)) for genetic variants in the region of interest for OPRM1 (top) and all-cause dementia (bottom). Right panels: Prior probabilities (top) and posterior probabilities (bottom) for colocalization hypotheses (H0–H4) calculated using a Bayesian framework. H0 indicates no association with either trait, H1 and H2 indicate associations with only one of the traits, H3 represents distinct causal variants for each trait, and H4 represents a shared causal variant. The dominance of H4 (yellow) in the green shaded region suggests strong evidence for colocalization, indicating a shared causal variant influencing both traits. The green shaded region highlights the genomic interval contributing most strongly to the H4 posterior probability.

#### C- Manhattan Plots Highlighting Association Signals for *OPRD1* and All-Cause Dementia on Chromosome 1

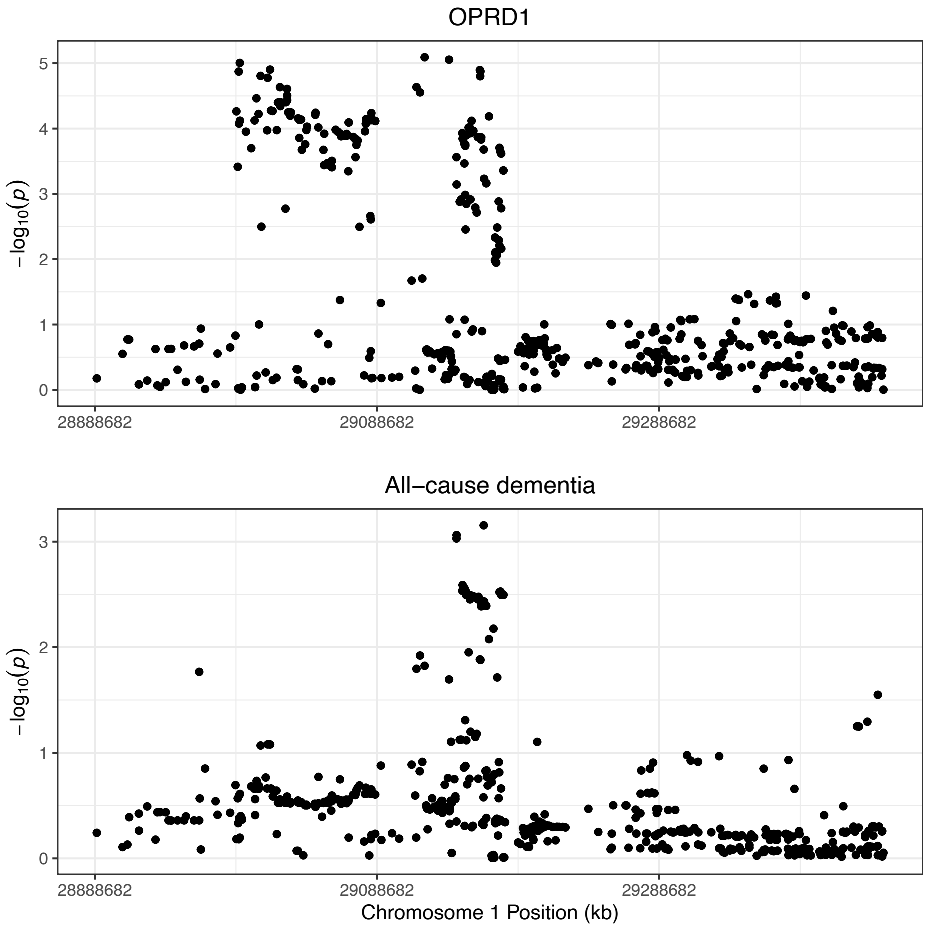


Manhattan plots showing association signals for *OPRD1* (top) and all-cause dementia (bottom) across Chromosome 1. Each point represents a genetic variant, with the y-axis indicating the -log10(p-value) of the association, and the x-axis representing genomic positions in kilobases (kb). Peaks in the plots highlight loci with stronger associations, suggesting potential regions of interest for exploring shared or distinct genetic influences on these traits.

#### D- Colocalization Analysis of *OPRD1* and All-Cause Dementia: Association Signals, Prior Assumptions, and Posterior Probabilities Across Hypotheses

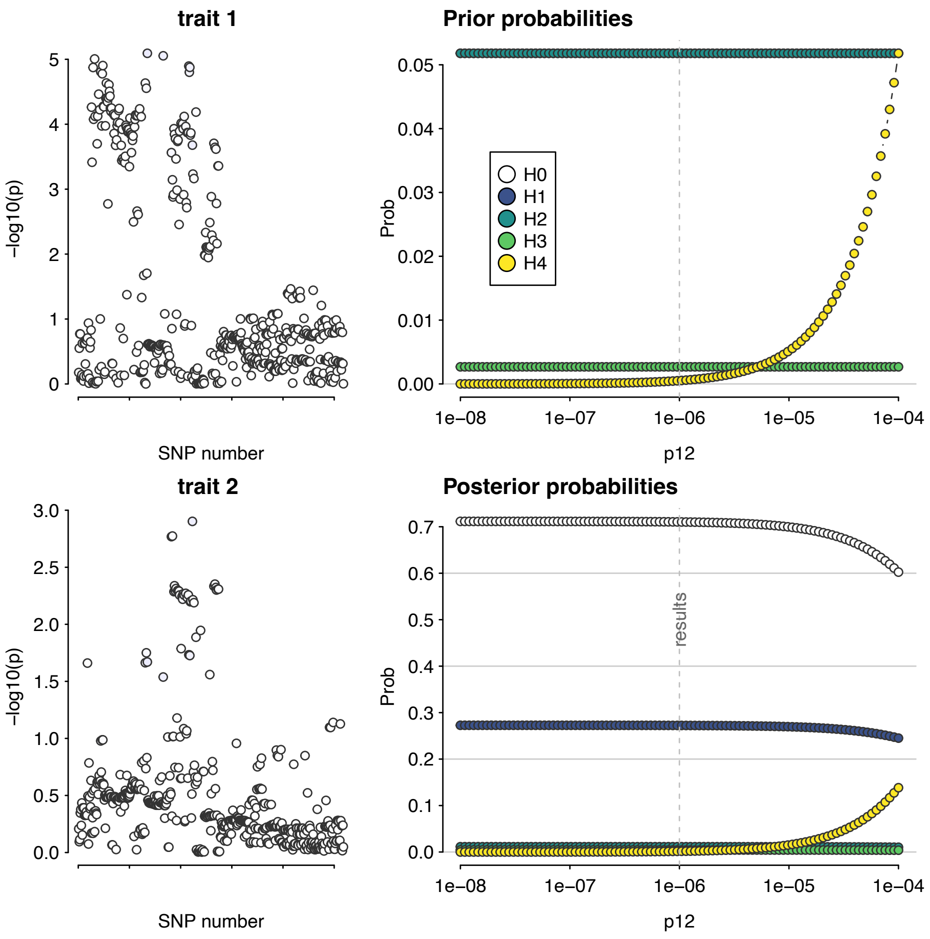


Colocalization analysis of *OPRD1* and all-cause dementia on Chromosome 1. Left panels: Manhattan plots displaying association signals (-log10(p-values)) for genetic variants in the region of interest for OPRD1 (top) and all-cause dementia (bottom). Right panels: Prior probabilities (top) and posterior probabilities (bottom) for colocalization hypotheses (H0–H4) calculated using a Bayesian framework. H0 represents no association, H1 and H2 represent associations with only one trait, H3 indicates distinct causal variants, and H4 suggests a shared causal variant. The prior probabilities (top right) illustrate initial assumptions, while the posterior probabilities (bottom right) reflect evidence from the observed data. The dominance of H0 in the posterior probabilities suggests limited evidence for association, and while the posterior for H4 (yellow) increases with higher prior probability (p12), it does not dominate, indicating weak support for a shared causal variant in this region.

### Supplemental Fig. 4

#### A- Scatterplot and Forest Plot of Genetically-Proxied μ-Opioid Receptor Perturbation and Alzheimer's disease risk

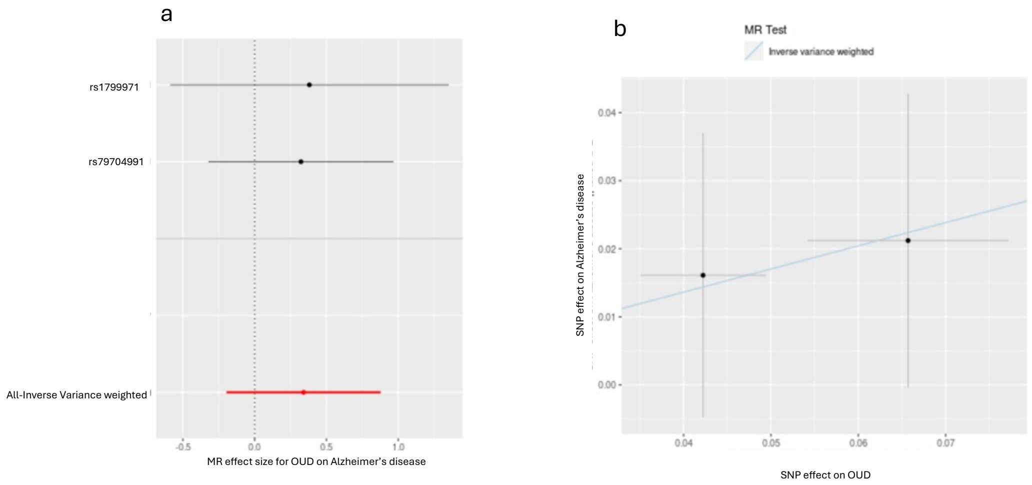


Cis-Mendelian randomization (MR) analysis of genetically-proxied μ-opioid receptor perturbation and Alzheimer's disease risk (Kunkle et al., 2019). (a) Scatter plot showing the effects of cis-acting SNPs on μ-opioid receptor perturbation (proxy for OUD) and Alzheimer's disease risk, with the slope of the regression line representing the causal effect estimated using the inverse variance weighted (IVW) method. (b) Forest plot displaying individual cis-acting SNP-specific causal estimates and the overall causal estimates. The results indicate no evidence of a causal relationship (IVW OR = 1.27 [0.87, 1.84]; p = 0.2).

#### B- Scatterplot and Forest Plot of Genetically-Proxied μ-Opioid Receptor Perturbation and Vascular dementia risk.

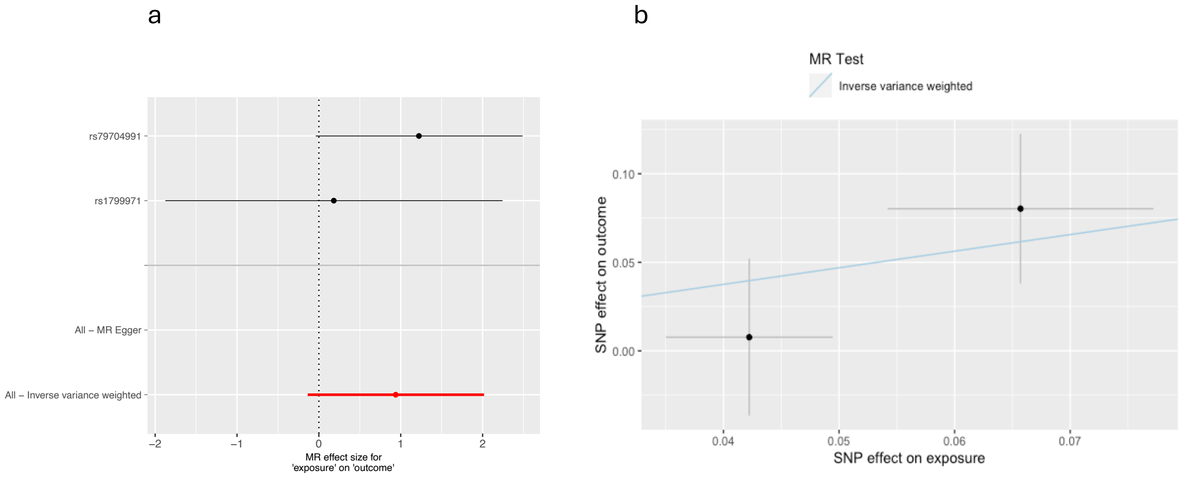


Cis-Mendelian randomization (MR) analysis of genetically-proxied μ-opioid receptor perturbation and vascular dementia risk (Fongang et al., 2024). (a) Scatter plot showing the effects of cis-acting SNPs on μ-opioid receptor perturbation and vascular dementia risk, with the slope of the regression line representing the causal effect estimated using the inverse variance weighted (IVW) method. (b) Forest plot displaying individual cis-acting SNP-specific causal estimates and the overall causal estimates. The results indicate no strong evidence of a causal relationship (IVW OR = 1.92 [0.91, 4.03]; p = 0.08).

#### C- Scatterplot and Forest Plot of Genetically-Proxied δ -Opioid Receptor Perturbation and Alzheimer’s disease risk.

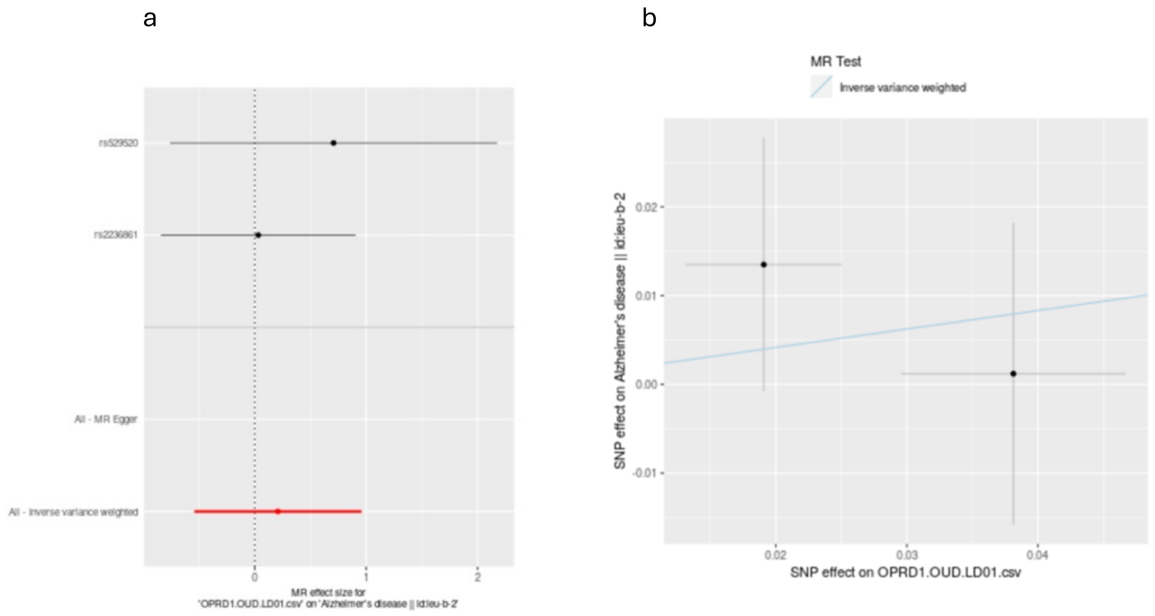


Cis-Mendelian randomization (MR) analysis of genetically-proxied δ-opioid receptor perturbation and Alzheimer’s disease risk (Kunkle et al., 2019). (a) Scatter plot showing the effects of cis-acting SNPs on δ-opioid receptor perturbation (proxy for OUD) and Alzheimer’s disease risk, with the slope of the regression line representing the causal effect estimated using the inverse variance weighted (IVW) method. (b) Forest plot displaying individual cis-acting SNP-specific causal estimates and the overall causal estimates. The results indicate no evidence of a causal relationship (IVW OR = 1.16 [0.69, 1.94]; p = 0.5).

#### D- Scatterplot and Forest Plot of Genetically-Proxied δ -Opioid Receptor Perturbation and Vascular dementia.

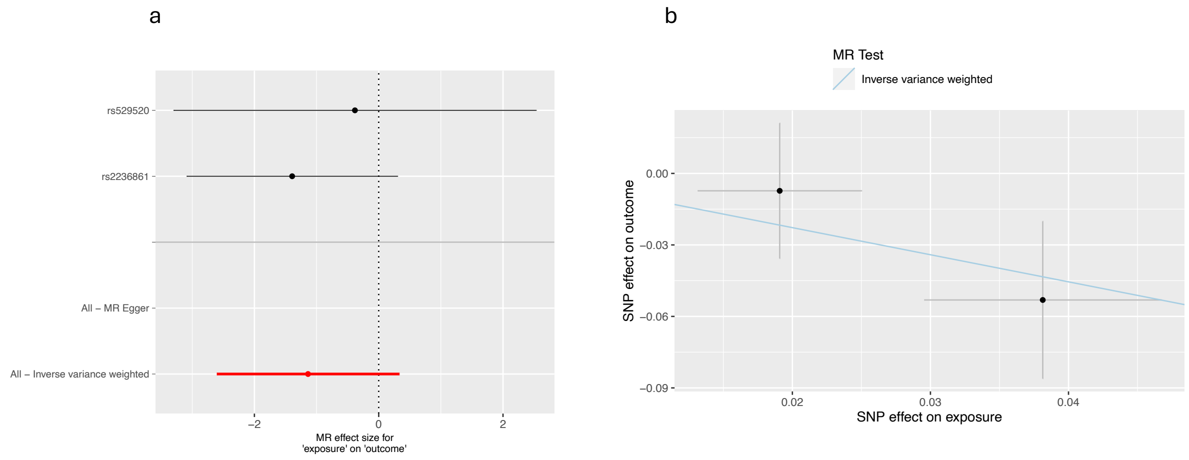


Cis-Mendelian randomization (MR) analysis of genetically-proxied δ-opioid receptor perturbation and vascular dementia risk (Fongang et al., 2024). (a) Scatter plot showing the effects of cis-acting SNPs on δ-opioid receptor perturbation (proxy for OUD) and vascular dementia risk, with the slope of the regression line representing the causal effect estimated using the inverse variance weighted (IVW) method. (b) Forest plot displaying individual cis-acting SNP-specific causal estimates and the overall causal estimates. The results indicate no strong evidence of a causal relationship (IVW OR = 0.45 [0.16, 1.26]; p = 0.1).

Supplemental Fig. 5

#### A- Scatterplot and Forest Plot of Genetic Association Between OUD and Ischemic Stroke in MR Analysis

a b

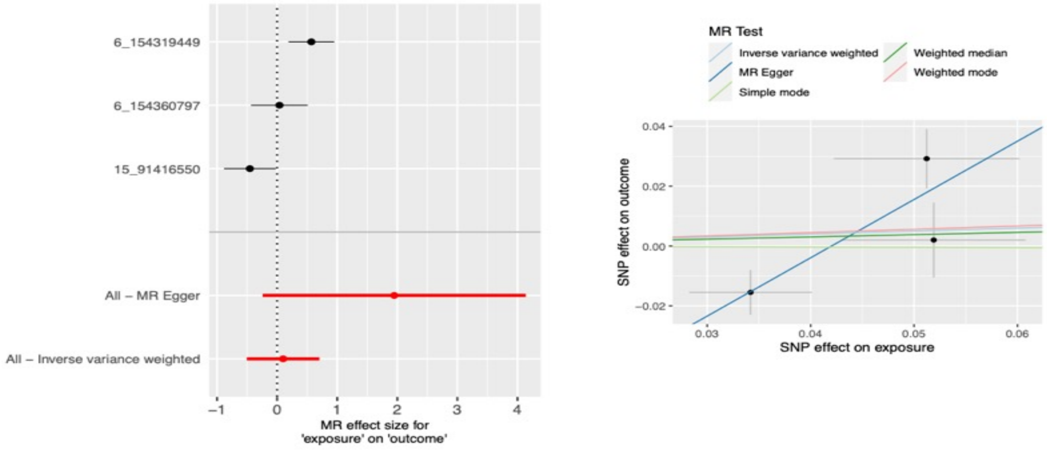


MR Analysis of the Association Between OUD and Ischemic Stroke Risk (Mishra et al., 2022). **a** Forest plot illustrating the MR effect sizes of OUD on ischemic stroke, using genome-wide significant (GWS) variants associated with OUD. **b** Scatter plot displaying SNP effects on OUD versus SNP effects on ischemic stroke, including estimates from multiple MR methods such as IVW, weighted median, MR-Egger, and weighted mode.

#### B- Scatterplot and Forest Plot of Genetic Association Between OUD and Large Artery Stroke in MR Analysis

a b

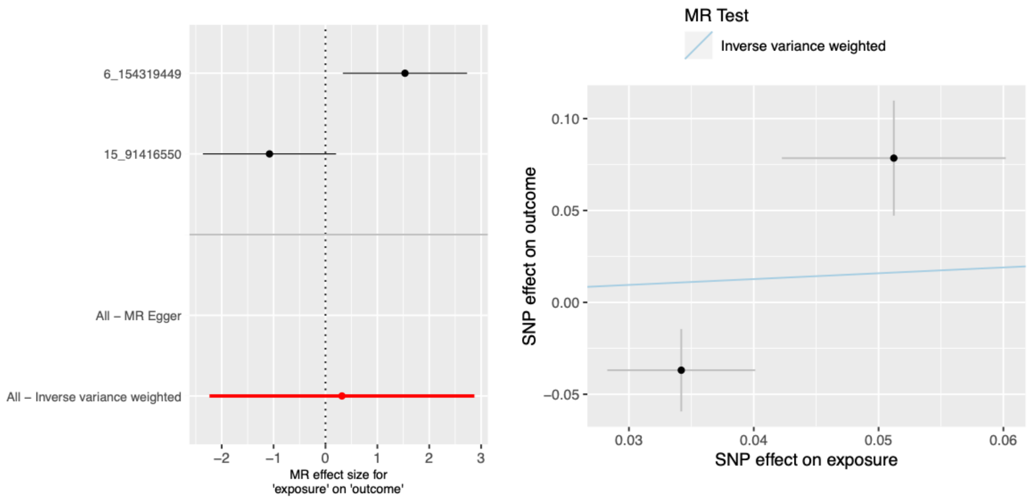


MR Analysis of the Association Between OUD and Large Artery Stroke Risk(Mishra et al., 2022). (a) Forest plot illustrating the MR effect sizes of OUD on large artery stroke, using genome-wide significant (GWS) variants associated with OUD. (b) Scatter plot showing SNP effects on OUD versus SNP effects on large artery stroke, with estimates derived using the inverse variance weighted (IVW) method.

#### C- Scatterplot and Forest Plot of Genetic Association Between OUD and Small Vessel Stroke in MR Analysis

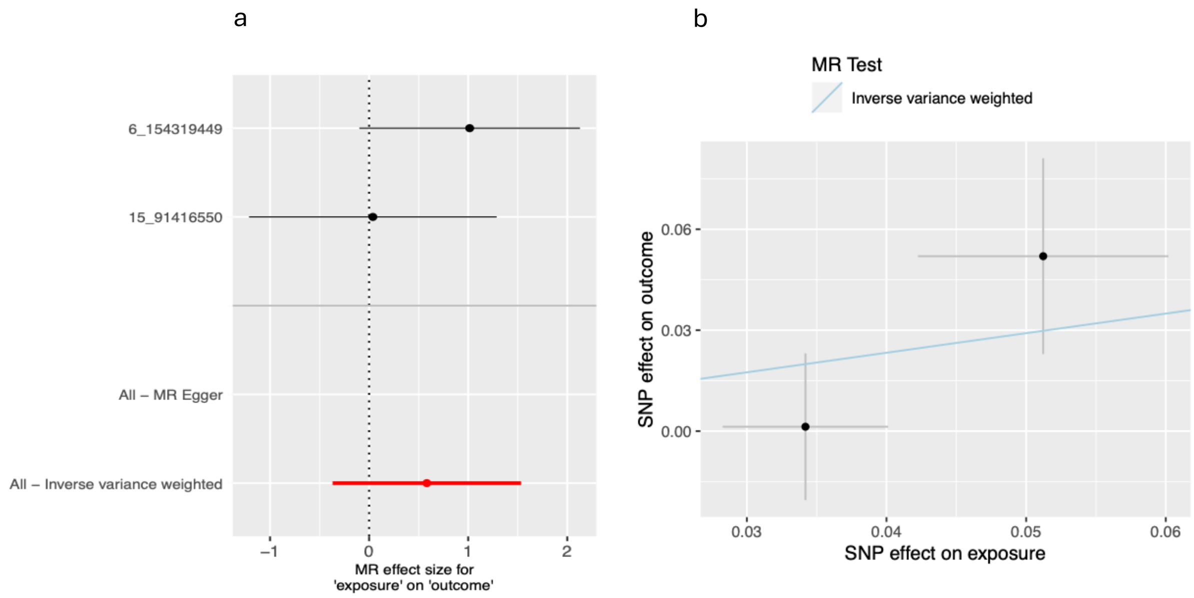


MR Analysis of the Association Between OUD and Small Vessel Stroke Risk(Mishra et al., 2022). (a) Forest plot illustrating the MR effect sizes of OUD on small vessel stroke, using genome-wide significant (GWS) variants associated with OUD. (b) Scatter plot showing SNP effects on OUD versus SNP effects on small vessel stroke, with estimates derived using the inverse variance weighted (IVW) method.

#### D- Scatterplot and Forest Plot of Genetic Association Between OUD and Cardioembolic Stroke in MR Analysis

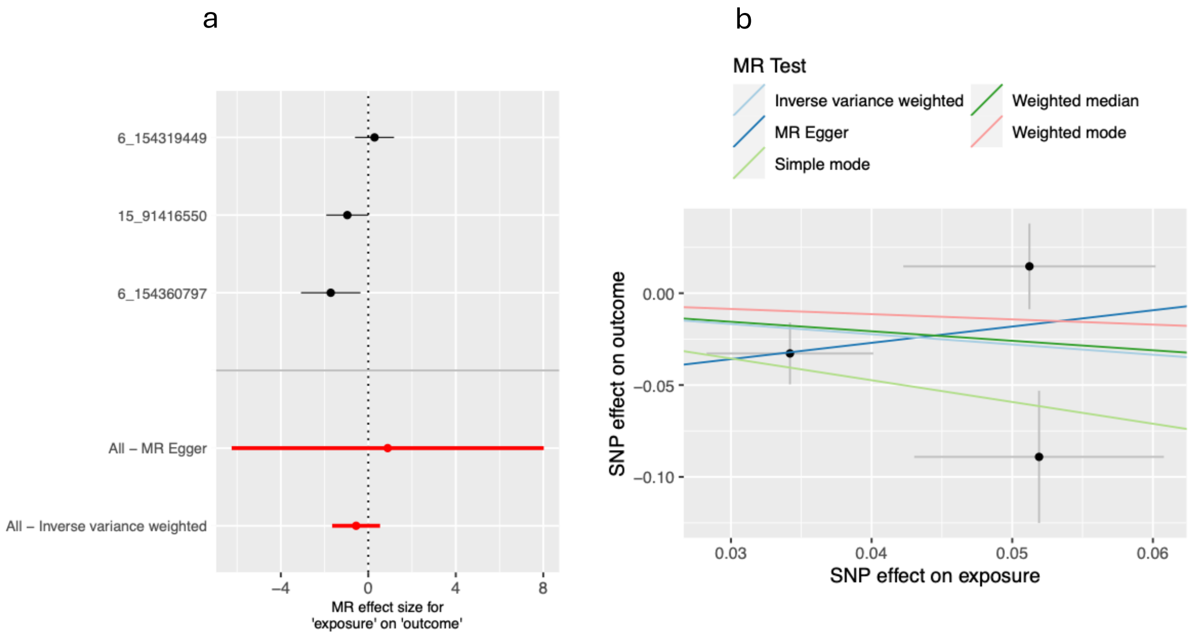


MR Analysis of the Association Between OUD and Cardioembolic Stroke Risk. (a) Forest plot illustrating the MR effect sizes of OUD on cardioembolic stroke, using genome-wide significant (GWS) variants associated with OUD. (b) Scatter plot showing SNP effects on OUD versus SNP effects on cardioembolic stroke, with estimates derived from multiple MR methods, including inverse variance weighted (IVW), weighted median, MR-Egger, weighted mode, and simple mode.

### Supplemental Fig. 6

#### Genetic Association between OUD and Stroke Subtypes.

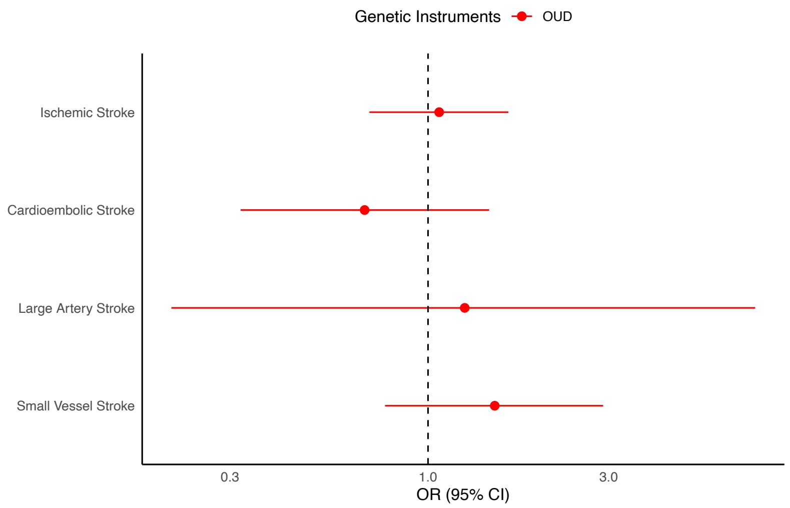


| OR(95%CI) | P |
|---|---|
| 0.70-1.63 | 0.74 |
| 0.32 - 1.45 | 0.31 |
| 0.21 - 7.31 | 0.80 |
| 0.77 – 2.90 | 0.22 |

Genetic associations between opioid use disorder and stroke subtypes. Estimates were generated using the inverse variance weighted method, are scaled per doubling of OUD prevalence.

### Supplemental Fig. 7

#### Genetic Association of OUD with Brain Structure Metrics in MR Analysis

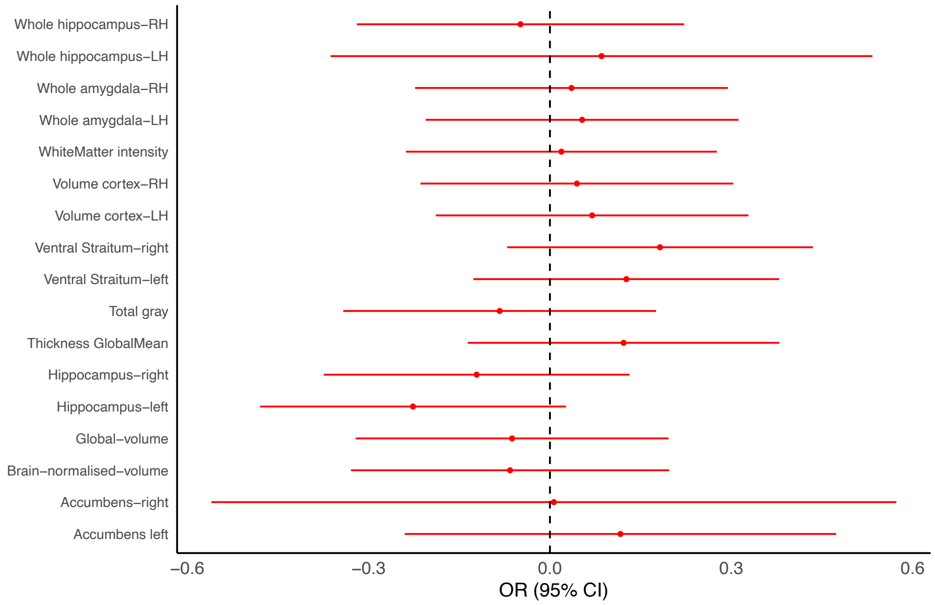


Forest plot showing the estimated effects (beta coefficients with 95% confidence intervals) of OUD (red) on various cross-sectional brain structure metrics, including hippocampus, amygdala, cortex, ventral striatum, accumbens, white matter intensity, total gray matter volume, and global mean thickness. The data for brain regions were obtained from the UK Biobank. The dashed vertical line represents the null hypothesis (no effect).

#### Supplemental Fig. 8

#### Mendelian Randomization Analysis of Opioid Use Disorder and Brain Structure: Forest and Scatter Plots of Genetic Associations

Total Gray

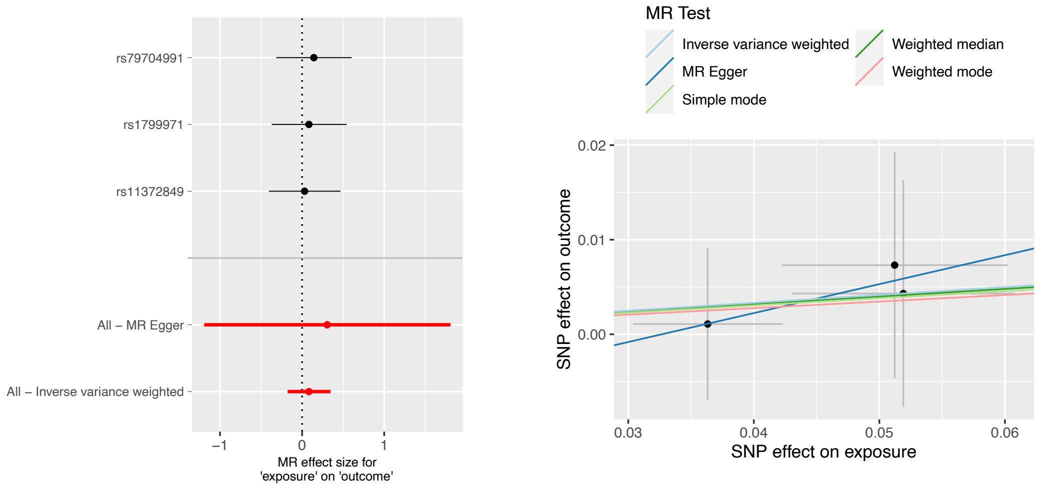


Amygdala-left hemisphere

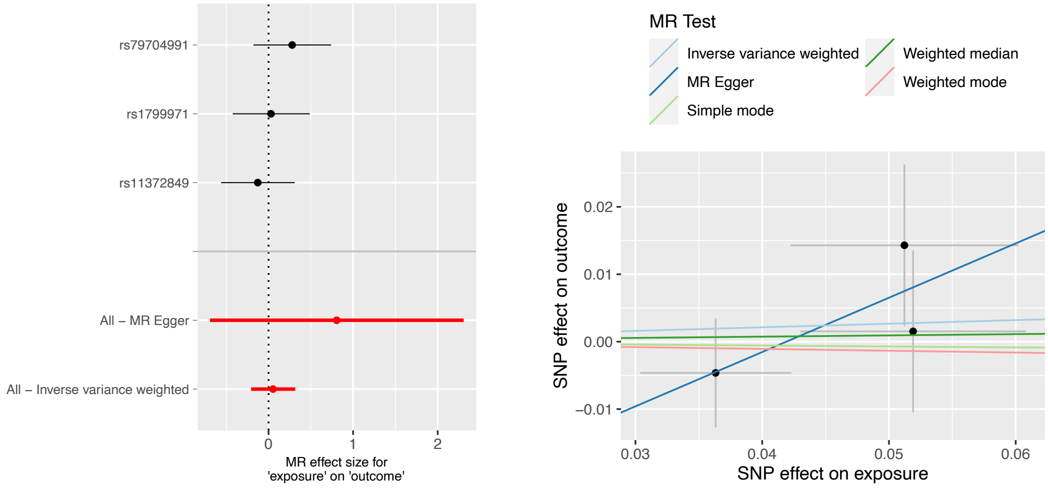

### Amygdala-right hemisphere

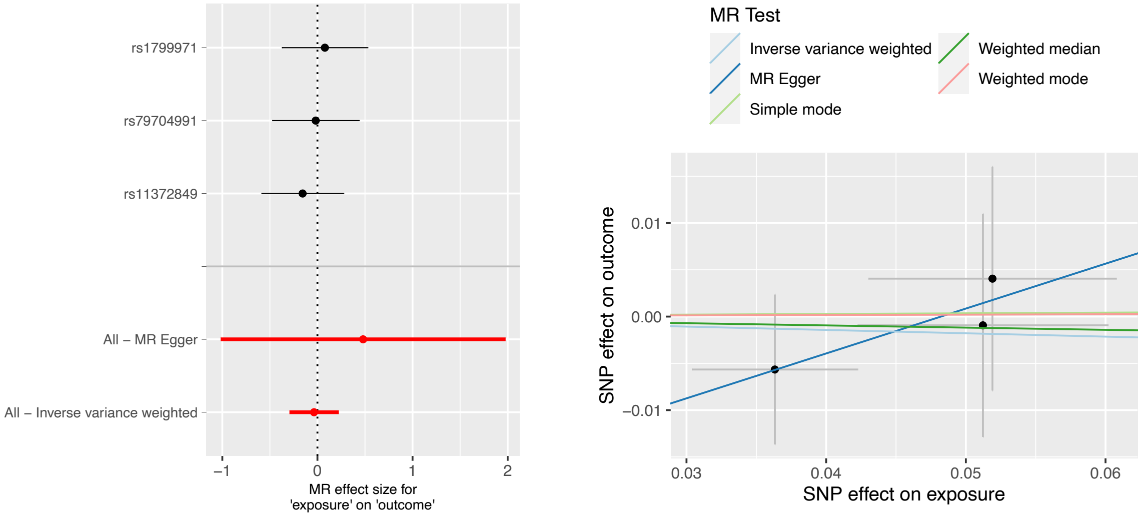


### Hippocampus left hemisphere

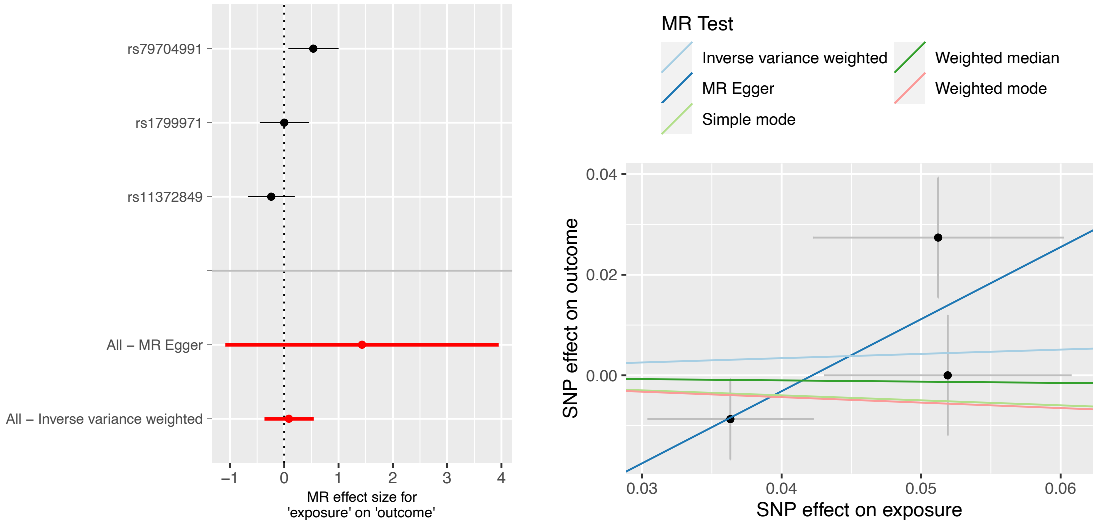

#### Hippocampus right hemisphere

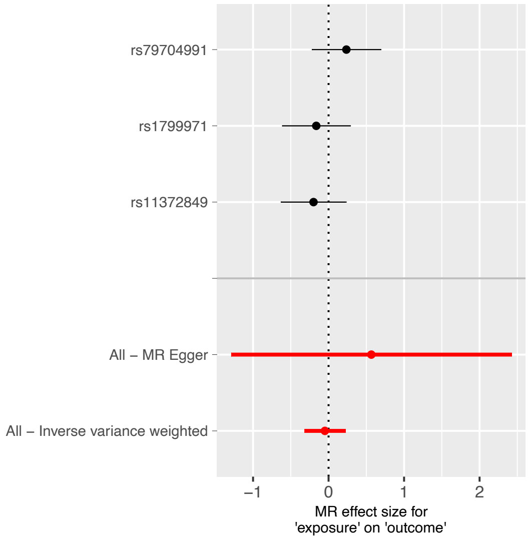


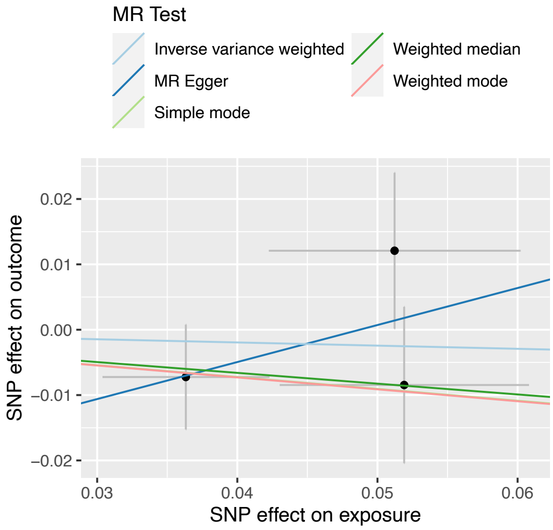


#### Cortex right hemisphere

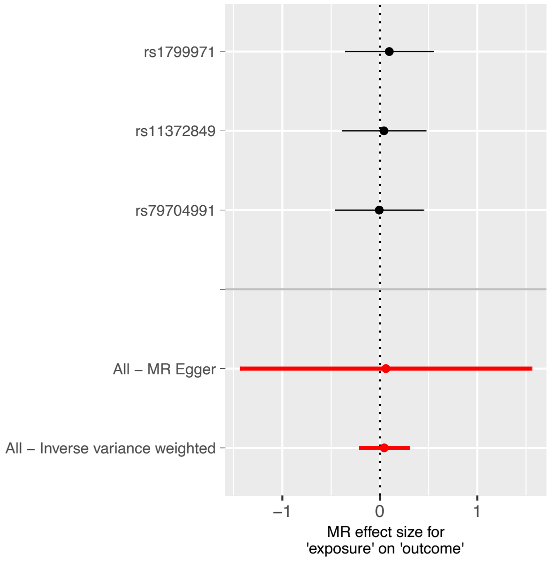


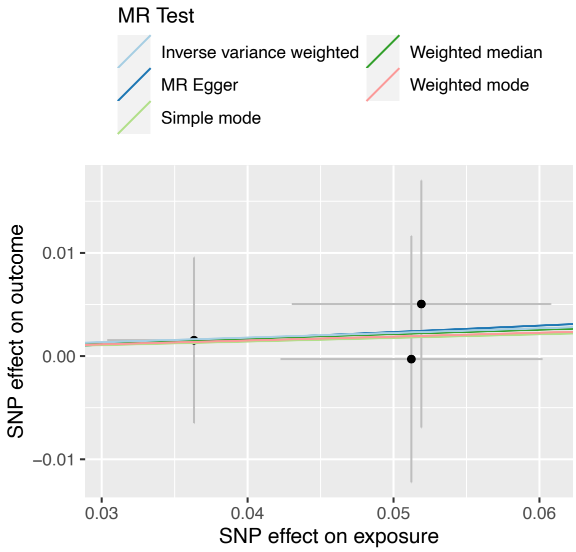

Cortex left hemisphere

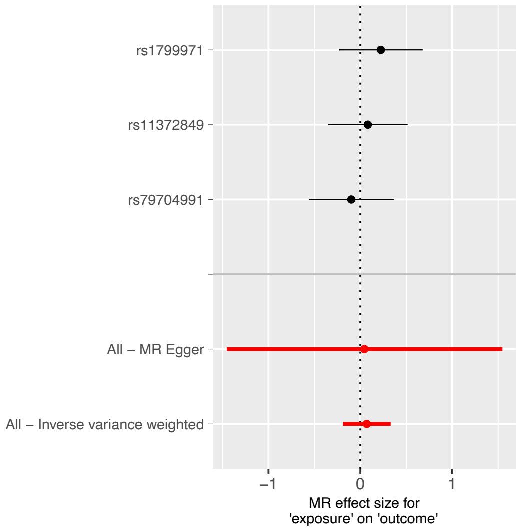


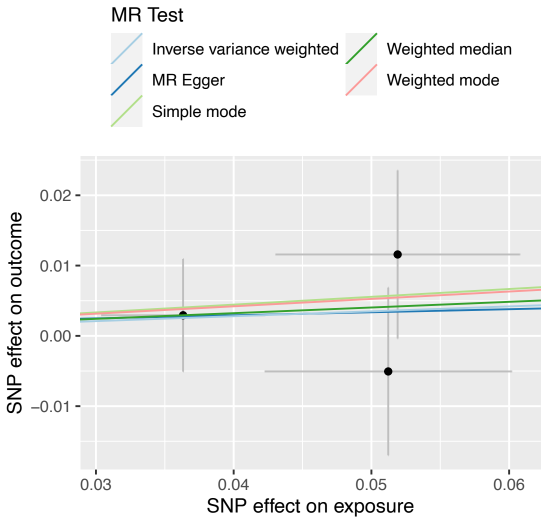


Accumbens left

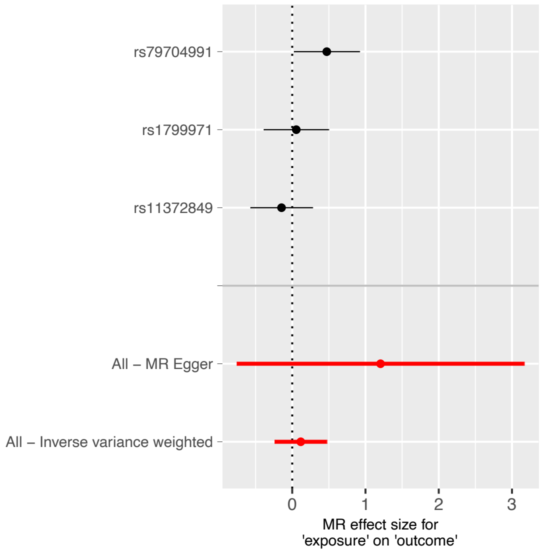


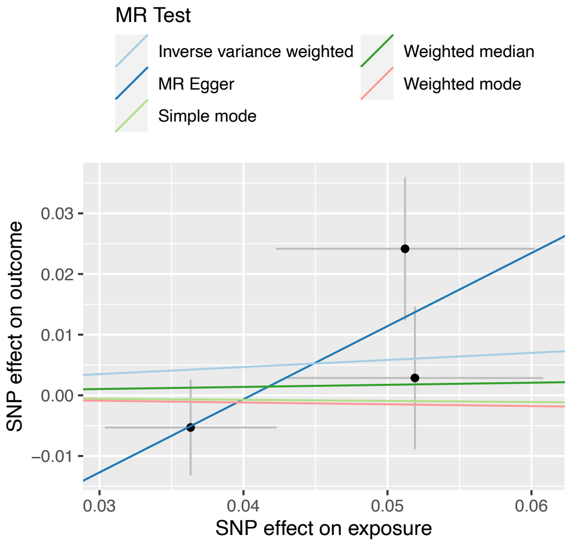

Accumbens right

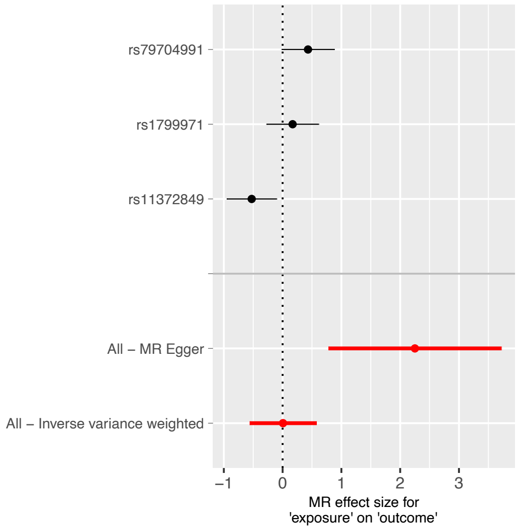


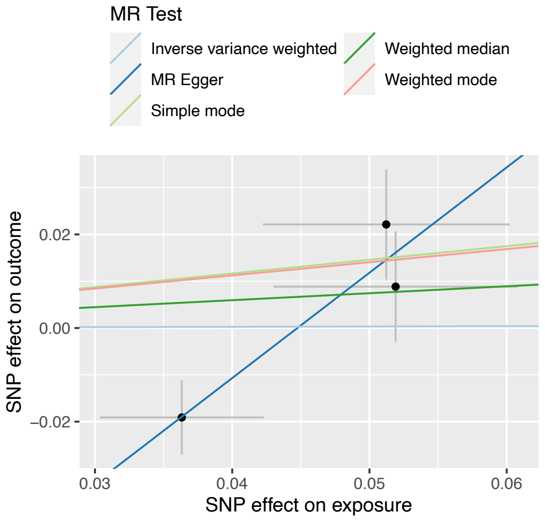


Ventral Striatum left

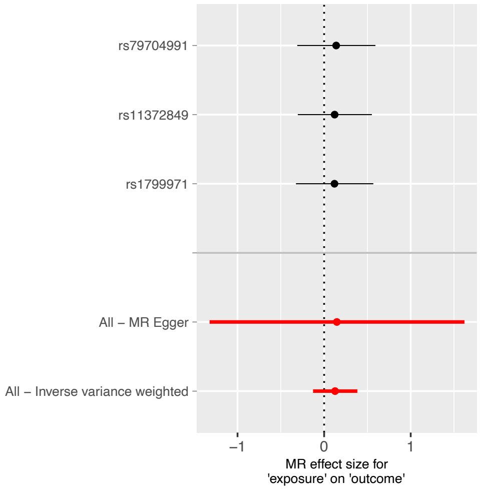


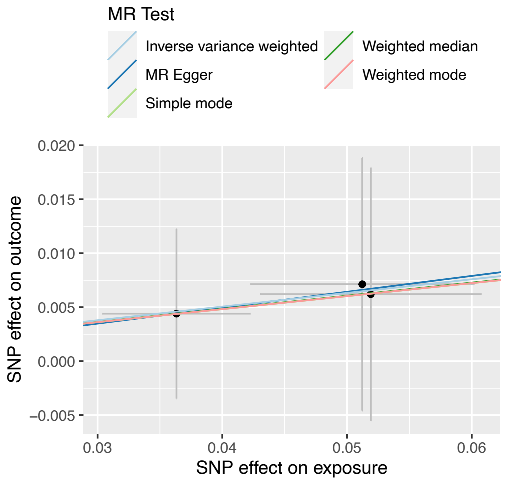

Ventral Striatum right

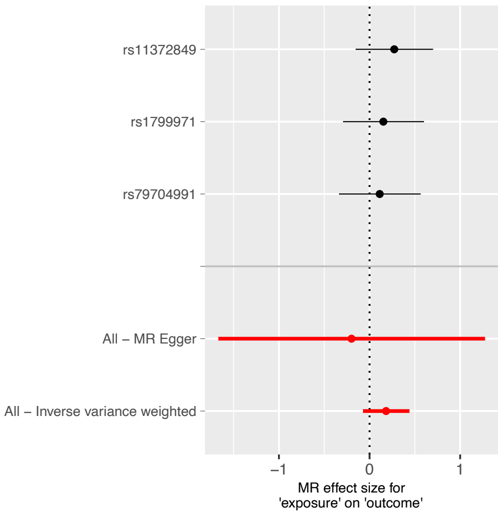


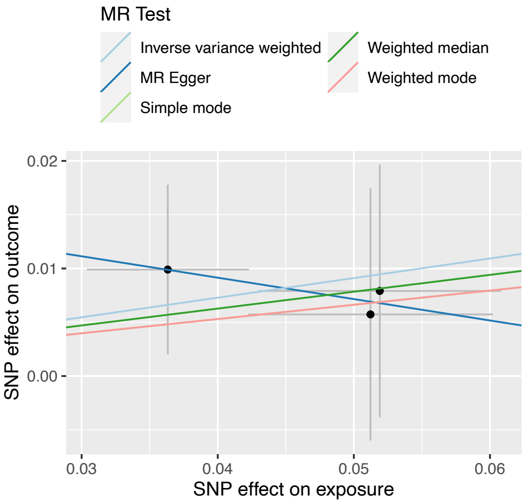


Hippocampus left volume

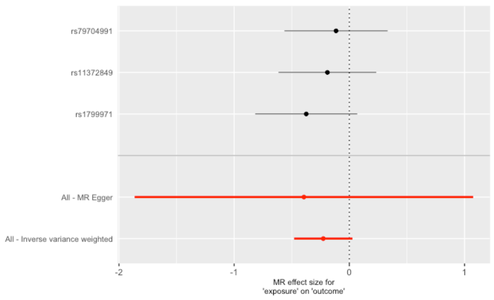


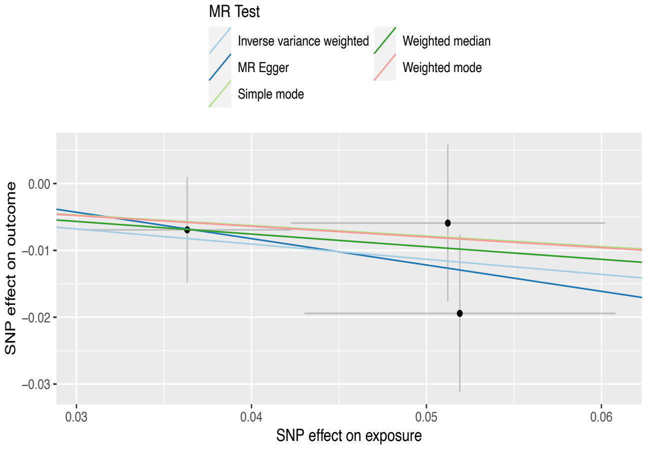

Hippocampus right volume

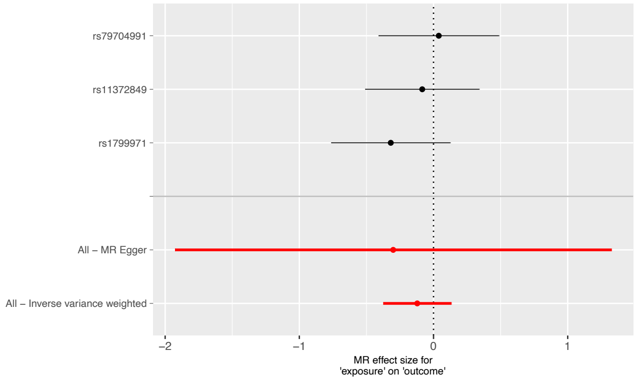


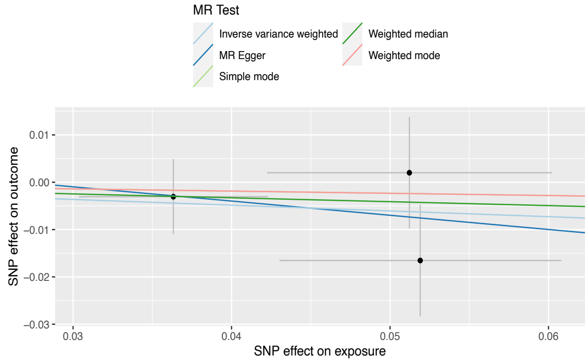


Brain Normalised volume

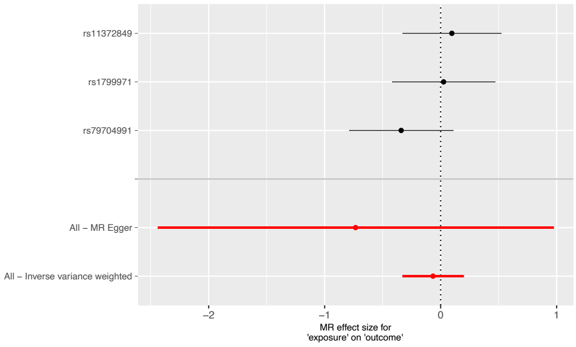


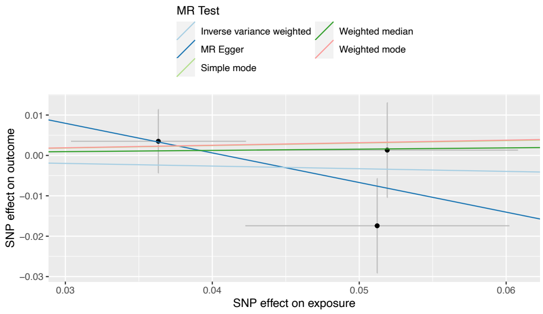


Global volume

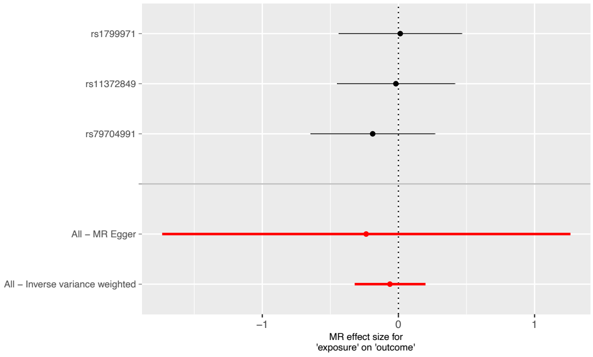


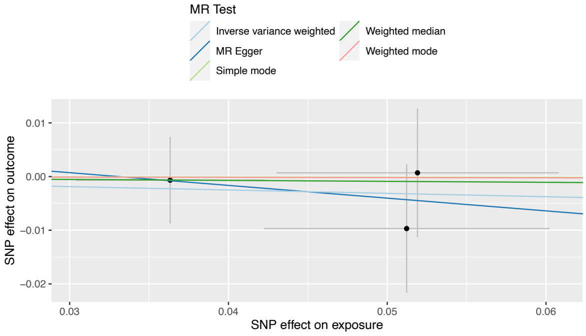

#### Thickness Global Mean

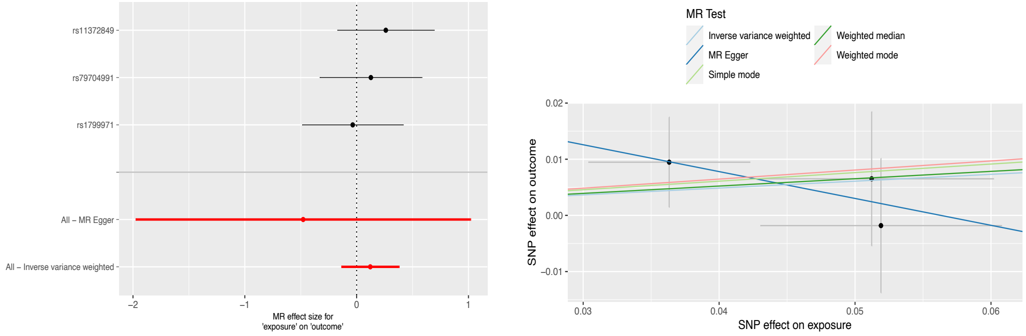


#### White Matter Hyperintensity

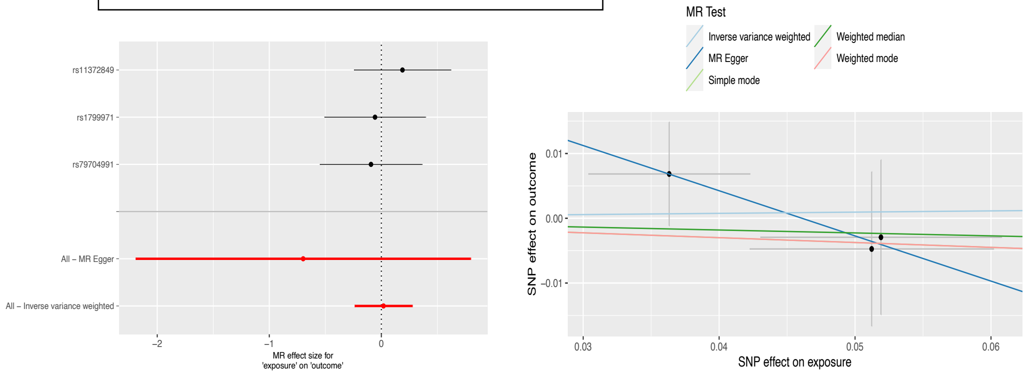


Mendelian Randomization (MR) analyses investigating the potential relationships between Opioid Use Disorder (OUD) and brain structural outcomes. The forest plots (left panels) display the MR effect sizes with corresponding 95% confidence intervals for individual SNPs and overall estimates using various MR methods, including Inverse Variance Weighted (IVW) and MR-Egger. The scatter plots (right panels) illustrate the SNP-specific associations between genetic effects on OUD and brain structure endophenotypes, with regression lines representing the different MR methods applied. These plots are provided for supplemental information and do not indicate evidence of causal relationships.
