## Supplementary Materials: Additional information supporting the main study. for "Opioid use disorder and brain health: observational and genetic associations"

### Supplemental.M1 MVP population for observational study

Recruitment was primarily conducted through invitational mailings and in-person enrolment at Department of Veterans Affairs (VA) facilities across the United States. Veterans who were active users of the Veterans Health Administration (VHA) and able to provide informed consent were eligible to participate. Data collection involved questionnaires, blood samples for genomic testing, and access to participants' VA electronic health records(Gaziano et al., 2016).

### Supplemental.M2 Polygenic Risk Score Study population

SNP pruning was performed to ensure independence, utilizing data from the European subpopulation in Phase 3 of the 1000 Genomes Project. The sample was obtained from the Center for Lifespan Changes in Brain and Cognition (LCBC) database at the Department of Psychology, University of Oslo, and data were collected using three different MRI scanners (Roe et al., 2024). Genetic ancestry factors (GAFs) were computed using established principal components methods. Importantly, all participants were healthy and reported no opioid abuse. This design allowed us to examine whether genetic propensity for opioid use was associated with longitudinal brain structural changes in healthy adults, independent of actual opioid exposure.

### Supplemental.M3 Covariate measurement

In the MVP, education was classified as a categorical variable based on self-reported levels: less than high school, high school diploma, some college credit, associate's degree, bachelor's degree, master's degree, and professional or doctoral degree. Household income was recorded as a categorical variable with the following ranges: less than $10,000, $10,000–19,999, $20,000–29,999, $30,000–39,999, $40,000–49,999, $50,000–59,999, $60,000–149,000, or $150,000 or more. Smoking status was categorized as daily, occasional, or not at all. Body mass index (BMI) was calculated using self-reported height and weight at enrolment. A history of head injury and post-traumatic stress disorder (PTSD) was also self-reported and coded as binary variables. Substance use disorders were defined based on a lifetime history of alcohol or cannabis dependence, identified by ICD codes in the electronic health records. Diabetes mellitus was reported in the enrolment survey. Mean systolic and diastolic blood pressure values were calculated using multiple measurements from the electronic health record. (Gaziano et al.,2015).
